# Clinical and Genomic Evaluation of 207 Genetic Myopathies in the Indian Subcontinent

**DOI:** 10.1101/2020.09.15.20193425

**Authors:** Samya Chakravorty, Babi Ramesh Reddy Nallamilli, Satish Khadilkar, Madhubala Singla, Ashish Bhutada, Rashna Dastur, Pradnya Gaitonde, Laura Rufibach, Logan Gloster, Madhuri Hegde

**Author notes:** Correspondence: *Samya Chakravorty Madhuri Hegde*, Lead Contact: Samya Chakravorty.

## Abstract

**Objective:** Inherited myopathies comprise more than 200 different individually rare disease-subtypes but when combined together have a high prevalence of 1 in 6000 individuals across the world. Our goal was to determine for the first time the clinical- and gene-variant spectrum of genetic myopathies in a substantial cohort study of the Indian subcontinent.

**Methods:** In this cohort-study, we performed the first large clinical exome sequencing (ES) study with phenotype correlation on 207 clinically well-characterized inherited myopathy-suspected patients from the Indian subcontinent with diverse ethnicities.

**Results:** Clinical-correlation driven definitive molecular diagnosis was established in 49% (101 cases; 95% CI, 42%-56%) of patients with the major contributing pathogenicity in either of three genes, *GNE* (28%; GNE-myopathy), *DYSF* (25%; Dysferlinopathy) and *CAPN3* (19%; Calpainopathy). We identified 65 variant alleles comprising 37 unique variants in these three major genes. 78% of the *DYSF* patients were homozygous for the detected pathogenic variant suggesting the need for carrier-testing for autosomal-recessive disorders like Dysferlinopathy that are common in India. We describe the observed clinical spectrum of myopathies including uncommon and rare subtypes in India: Sarcoglycanopathies *(SGCA/B/D/G)*, Collagenopathy *(COL6A1/2/3)*, Anoctaminopathy *(ANO5)*, telethoninopathy *(TCAP)*, Pompe-disease *(GAA)*, Myoadenylate-deaminase-deficiency-myopathy *(AMPD1)*, myotilinopathy *(MYOT)*, laminopathy *(LMNA)*, HSP40-proteinopathy *(DNAJB6)*, Emery-Dreifuss-muscular-dystrophy *(EMD)*, Filaminopathy *(FLNC)*, TRIM32-proteinopathy *(TRIM32)*, POMT1-proteinopathy *(POMT1)*, and Merosin-deficiency-congenital-muscular-dystrophy-type-1 *(LAMA2)*. 13 Patients harbored pathogenic variants in >1 gene and had unusual clinical features suggesting a possible role of synergistic-heterozygosity / digenic-contribution to disease presentation and progression.

**Conclusions:** Application of clinically-correlated ES to myopathy diagnosis has improved our understanding of the clinical and genetic spectrum of different subtypes and their overlaps in Indian patients. This, in turn, will enhance the global gene-variant-disease databases by including data from developing countries/continents for more efficient clinically-driven molecular diagnostics.

## INTRODUCTION

To date, >250 genes are associated with various inherited neuromuscular disorders (NMDs) (1-4) comprising a broad heterogeneous group of genetic myopathies. However, significant numbers of affected patients remain without a definitive molecular diagnosis due to novel gene or multiple gene associations with disease (5, 6). The autosomal-dominant forms represent <10% of all genetic myopathies (6-8) and are usually associated with milder clinical-presentation and adult-onset. Whereas, for the more common autosomal-recessive forms, multiple monogenic subtypes have been identified including those that make up the limb-girdle muscular dystrophies (LGMDs) (5, 9). Definitive molecular diagnosis is a prerequisite for patient participation in clinical-trials or precision medicine treatment and management. Hence, it is important to understand the genetic etiologies and subtype prevalence in different populations to enhance global variant databases for more efficient and accurate gene-variant classification and diagnosis.

Several studies reported the use of next-generation-sequencing (NGS) for molecular diagnosis in small/large genetic myopathy patient-cohorts in different countries (6, 10-12). But the application of exome sequencing (ES) for a large cohort of myopathy patients from the Indian subcontinent has not been reported previously. Similar to the rising awareness of the importance of African genomes or exomes in global public health and enhancing public databases, it is important to study the populations of the Indian subcontinent and Asia as a whole comprising of the largest proportion of the world population which offers diverse ethnic and genetic backgrounds (13-16). Moreover, the sociocultural belief systems and traditions have led to community marriages, consanguinity and intra-communal exogamy in sections of the Indian population. Here, we performed the first large-scale ES study on 207 clinically well-characterized inherited myopathy-suspected patients of diverse ethnicities from the Indian subcontinent to identify the causative gene and disease-subtypes. Our results yield interesting observations on Indian myopathy clinical and gene-variant landscape and approaches to both clinical and genetic diagnosis in muscular dystrophy and myopathy in general.

## MATERIALS AND METHODS

### Standard Protocol Approvals, Registrations, and Patient Consents

Written, informed consent was obtained from all individuals or minors’ parent or legal guardian or next of kin for the publication of any potentially identifiable images or data included in this article. Written informed consents were obtained from all participants of the study for all procedures according to approved Ethics committee Institutional Review Board protocol. All participants (including parents or legal guardians in case of minor patients) were provided pre- and post-test routine genetic counseling.

### Patients and Study Design

In an effort to understand the genetic basis of myopathies, over a 3-year period (2016-2019) a total of 207 patients with clinical suspicion of an inherited myopathy were recruited (see Questionnaires in Supplementary Material) based on the following inclusion and exclusion criteria. Inclusion criteria: Patients with a suspected clinical diagnosis of genetic myopathy. Exclusion Criteria: 1. Patients with confirmed diagnosis of Duchenne muscular dystrophy (DMD), facioscapulohumeral muscular dystrophy (FSHD), myotonic dystrophy (DM1 and DM2), mitochondrial myopathy and Acquired myopathies. DMD, myotonic dystrophies and FSHD were genetically diagnosed. Mitochondrial myopathies were diagnosed based on muscle biopsy immunohistochemistry and/or genetic diagnosis. We used modified Gomori trichrome, cytochrome oxidase (COX) and nicotinamide adenine dinucleotide (NADH) stains for the demonstration of mitochondrial dysfunction, supported by electron microscopy as needed. There was no age restriction and the patients were recruited from across the Indian subcontinent at a single tertiary care hospital.

### Clinical Evaluation

All patients underwent comprehensive clinical evaluation. Clinical details including age of onset, initial symptoms, region in which weakness started first, and functional status of the patient was collected whenever available for all patients recruited in the study. Pattern of weakness including upper limbs or lower limbs, or both, proximal or distal or both, symmetrical or asymmetrical, and differential weakness, was determined. Detailed pedigree analysis was performed. Available affected family members were examined. Detailed history regarding the origins and ancestry and marriage customs practiced in the specific ethnic community was documented. Patients underwent detailed clinical examination with focus on the motor system. Hypertrophy, atrophy of muscles was determined from inspection. Neck flexors, extensors and other total 16 pairs of muscles (trapezius, deltoid, serratus anterior, biceps, brachioradialis, triceps, wrist flexors and extensors, ileopsoas, hip adductors, abductors, extensors, quadriceps, hamstrings, tibialis anterior, gastrocnemius-soleus) were examined as per the MRC grading system of 0 to 5. Small muscles of hands, feet, and paraspinal muscle were also examined. Serum creatine kinase was determined at the time of presentation prior to electromyography. Electromyography and nerve conduction studies were performed in all patients. Electrocardiography was done in all patients and Echocardiography was done in selected symptomatic patients.

### Muscle Biopsy Procedure

Muscle biopsy procedure was performed for only the consenting patients as per routine methodology. Muscle biopsy was performed from clinically affected muscle with power between MRC grades 3 to 4. Biopsy specimens were snap frozen in chilled isopentane for frozen sections and later taken in formalin for paraffin embedding. Routine assessment of morphological changes was carried out on frozen and paraffin sections. Biopsy specimens from those patients who consented for the procedure were grouped into dystrophic, myopathic, dystrophic with inflammation, myositis, mixed (neurogenic and myopathic) changes, and some no diagnosis. Histochemical studies were done using ATPase and NADH-TR (nicotinamide adenine dinucleotide (NAD) + hydrogen (H) and tetrazolium reductase) stains. Frozen sections of 6 microns were stained with dystrophin 1 (C terminus-mouse monoclonal Dy8\6C5), dystrophin 2 (mid-rod domain mouse monoclonal Dy4\6D3), dysferlin (mouse monoclonal NCL-Hamlet-2), and α (Ad1\20A6), β (Bsarc\5B1), γ (35 DAG\21B5) and δ (Dsarc 3\12C1) sarcoglycan antibodies (Novocastra labs), using peroxidase method and standard controls.

### Molecular Diagnosis by Exome Sequencing

Exome sequencing was performed in a Clinical Laboratory Improvement Amendments and College of American Pathologists (CLIA-CAP)-certified laboratory. Peripheral intravenous blood was collected from enrolled patients into 10-ml commercial tubes containing ethylene-diamine tetraacetic acid (EDTA). These samples were shipped immediately overnight on ice to our laboratory for further analysis. DNA was extracted from whole blood using the Qiagen genomic DNA extraction Kit according to the manufacturer’s instructions. We performed exome sequencing using Agilent V5Plus exome capture kit to identify disease causative variants. Sequencing was performed using the Illumina HiSeq 2500 sequencing system with 100-basepair (bp) paired-end reads (similar to bidirectional Sanger sequencing), with an average coverage of 100X in the target region. The target region includes the exon and 10 bp of flanking intronic region. NextGENe software mapped the DNA sequence reads to the published human genome build UCSC hg19 reference sequence. Primary data analysis was also performed using Illumina DRAGEN Bio-IT Platform v.2.03. Secondary and tertiary data analysis was also performed using PerkinElmer’s internal ODIN v.1.01 software for SNVs and Biodiscovery’s NxClinical v.4.3 or Illumina DRAGEN Bio-IT Platform v.2.03 for copy number variant (CNV) and absence of heterozygosity (AOH). Reads were aligned and filtered as described previously (17, 18). In short, in the filtering process, variants in genes that are not relevant to the patient’s clinical phenotypes were filtered in the clinical setting using population-wide minor allele frequency, predicted effect on protein function or splicing, literature evidence, and disease-variant databases such as the Human Gene Mutation Database (http://www.hgmd.cf.ac.uk), ClinVar (http://www.clinvar.com), Online Mendelian Inheritance of Man (https://www.ncbi.nlm.nih.gov/omim), genome Aggregation Database (gnomAD: https://gnomad.broadinstitute.org/), and using our internal Emory Genetics Lab (EGL) EmVClass database (http://www.egl-eurofins.com/emvclass/emvclass.php). Variants that do not meet QC metrics, such as those with poor coverage (<20×), were considered less likely to be real, treated as false positives, and therefore filtered. Variants with a minor allele frequency of >0.01 are polymorphisms by definition and less likely to be pathogenic. Silent changes and intronic variants beyond the consensus splice donor/acceptor sequences or beyond the 10bp intronic flanking sequences are less likely to be pathogenic and were filtered in the analysis unless a known clinically significant rare deep-intronic variant is identified. Rare single nucleotide variants (SNVs) meeting internal quality assessment guidelines are confirmed by Sanger sequence analysis.

Molecular diagnosis was performed by classifying variants with clinical data correlation to identify most likely causal disease gene and as per American College of Medical Genetics and Genomics (ACMG) guidelines (19) and with clinical data correlation. Diagnostic yield was calculated based on definitive diagnosis of patients harboring pathogenic or likely pathogenic variants in autosome-recessive/dominant, and X-linked myopathy genes. Prevalence of each identified myopathy subtype identified by our clinical exome sequencing diagnostic program was established and compared. When rare reportable variants in our study were not found in NIH ClinVar (https://www.ncbi.nlm.nih.gov/clinvar/), nature of variant, literature evidence and correlation with established specific clinical symptoms of the patient’s myopathy was used to identify most likely causal gene and to classify the variant, keeping in mind ACMG guidelines. When no literature evidence was found, the variant was classified based on nature of variant such as nonsense variant or frameshift variant causing a premature stop codon or a stop-loss or a large deletion or duplication in genes known to affect by loss of function modality, as well as correlation with established specific clinical symptoms of the patient’s myopathy. High-confidence rare variants were taken as novel when not found in public databases and included in the analysis to interpret relevant disease type causality with clinical correlation. Patient cases with variants of uncertain significances (VUSs) were not included in the molecular diagnostic yield calculations.

## RESULTS

### Molecular diagnosis of recruited patients

A total of 207 unrelated myopathy-suspected patients were recruited in the current study from diverse regions of India with different ethnicities and religion, as well as varied social and marriage customs. Molecular diagnosis was established in 49% (101/207; 95% CI, 42%-56%) of the patients with the majority having pathogenic variants identified in one of the following genes, *GNE* (31%; 31/101), *DYSF* (27%; 27/101), or *CAPN3* (19%; 19/101), indicating that these genes are the major contributors to genetic myopathy in India (Figure 1). Autosomal-recessive forms were much more frequent constituting 95/101 (94%) of the genetically-diagnosed patients. Uncommon autosomal/X-linked recessive-subtypes identified were: 5 sarcoglycanopathies (LGMD R3-R6; 5%), 3 anoctaminopathies (LGMD R12 anoctamin-related; 3%), 2 myoadenylate-deaminase-deficiency-myopathies (2%), 2 Pompe disease (2%), 2 telethoninopathies (LGMDR7 telethonin-related; 2%), 1 X-linked Emery-Dreifuss-muscular-dystrophy (EDMD1 emerin-related; 1%), 1 Merosin-deficiency-congenital-muscular-dystrophy-type 1 (MDC1 or LGMDR23 laminin α2-related; 1%), 1 POMT1-proteinopathy (LGMDR11 POMT1-related; 1%). Autosomal-dominant-subtypes were less common with 3 laminopathies (EDMD2; 3%), 1 myotilinopathy (Myofibrillar Myopathy; 1%), and 1 HSP40-proteinopathy (LGMDD1 DNAJB6-related; 1%). 14 patients had no reportable variants and were truly negative for this study.

**Figure 1.**
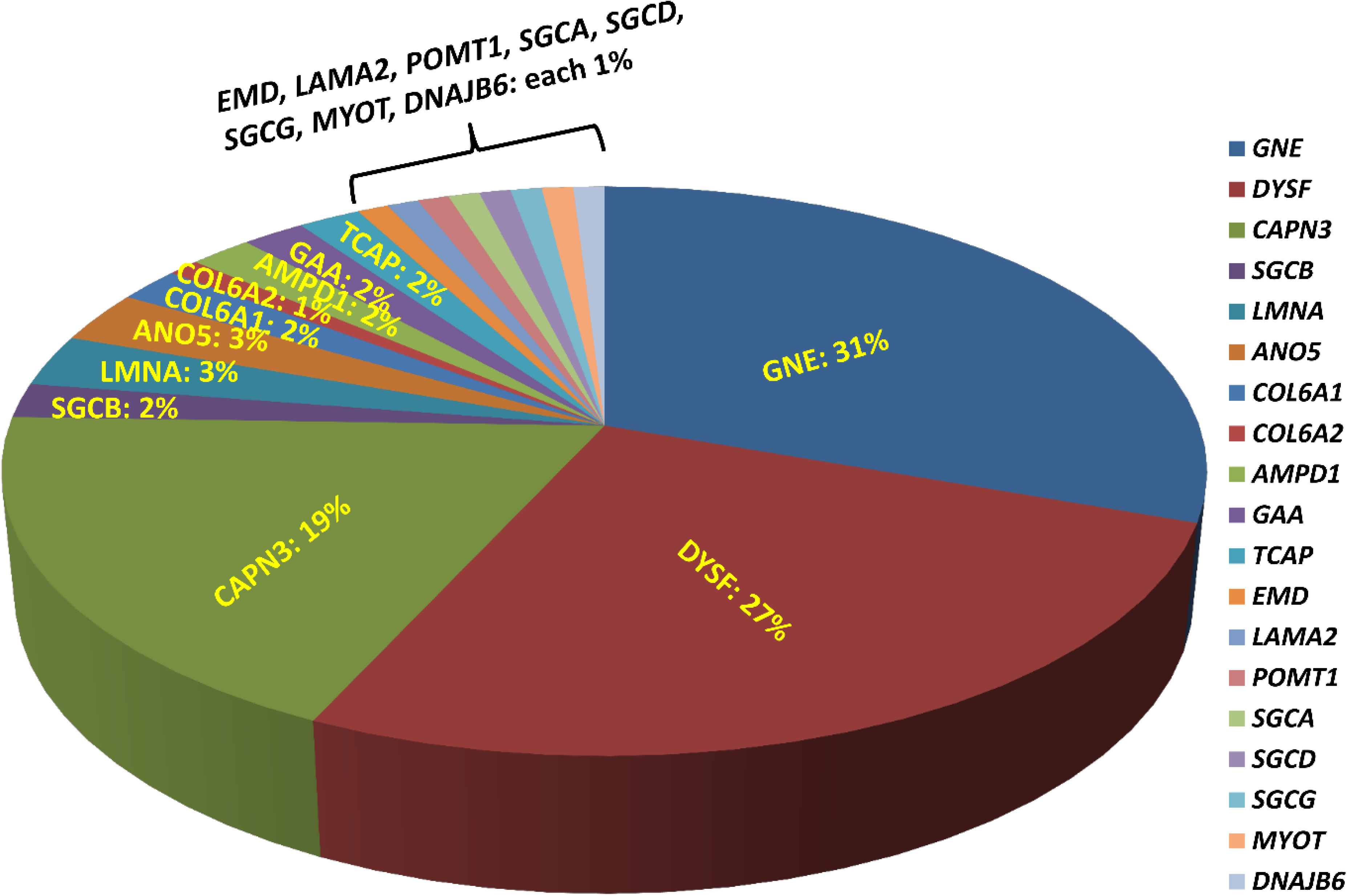
Molecular diagnosis with different genetic myopathy subtypes. Molecular diagnosis using exome sequencing has been established in 49% of the patients with the majority having pathogenic variants identified in one of the following genes *GNE* (31%), *DYSF* (27%), *CAPN3* (19%), indicating that these genes are likely the major contributors to genetic myopathy-like phenotype.

We identified 65 variant alleles comprised of 37 unique variants in the three major genes *(GNE, DYSF, CAPN3)*. A total of 14 unique variants were identified in *GNE* (Figure 2A). The present study confirmed the absence of a mutational hot-spot region in *GNE* in the Indian patients. The majority of the variants detected in *GNE* (12) were missense variants located throughout the *GNE* gene, suggesting an important role of missense changes in causing GNE-myopathy in people of Indian subcontinent origin (Table 1). The most common pathogenic variants c.2179G>A (p/V727M) and c.1760T>C/c.1853T>C (p.I587T/p.I618T) were detected in most of the *GNE* patients (Figure 2D) with extensive homozygosity observed for the c.1760T>C (p.I587T) common pathogenic variant. Only one nonsense variant c.385C>T (p.R129X), one frameshift variant c.397_398dupAT and no splice site variants were identified in *GNE*.

**Table 1.**
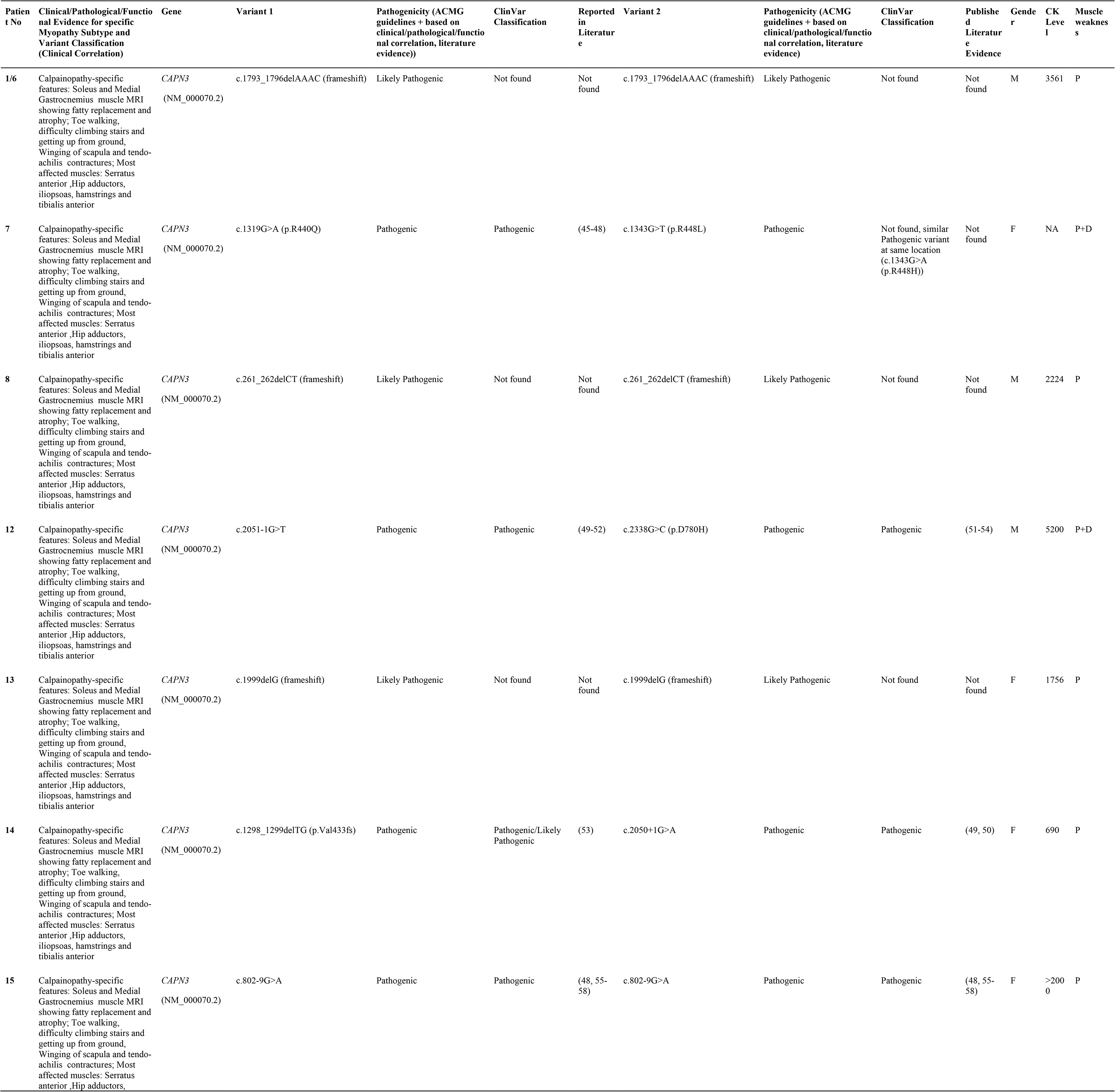
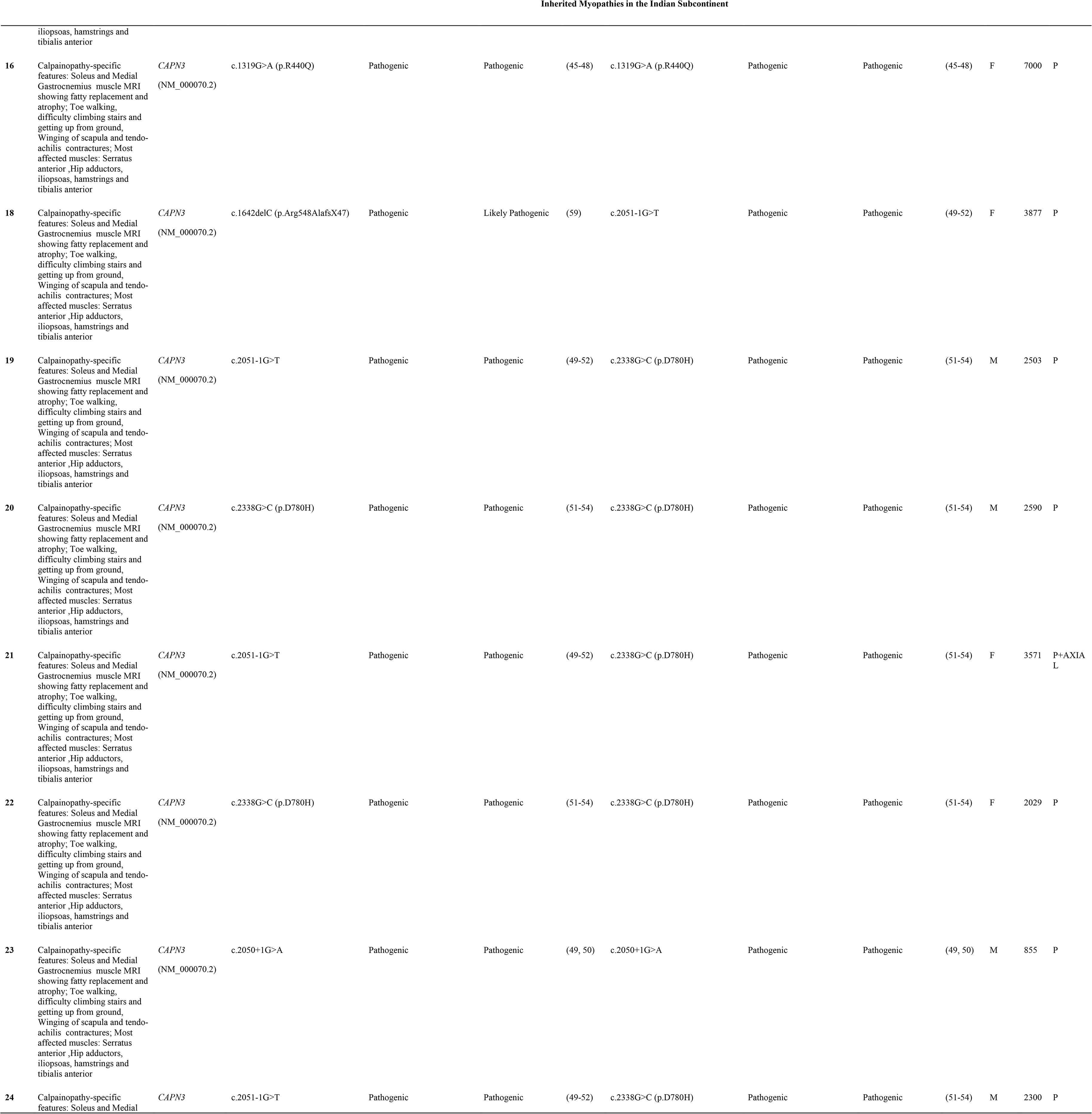
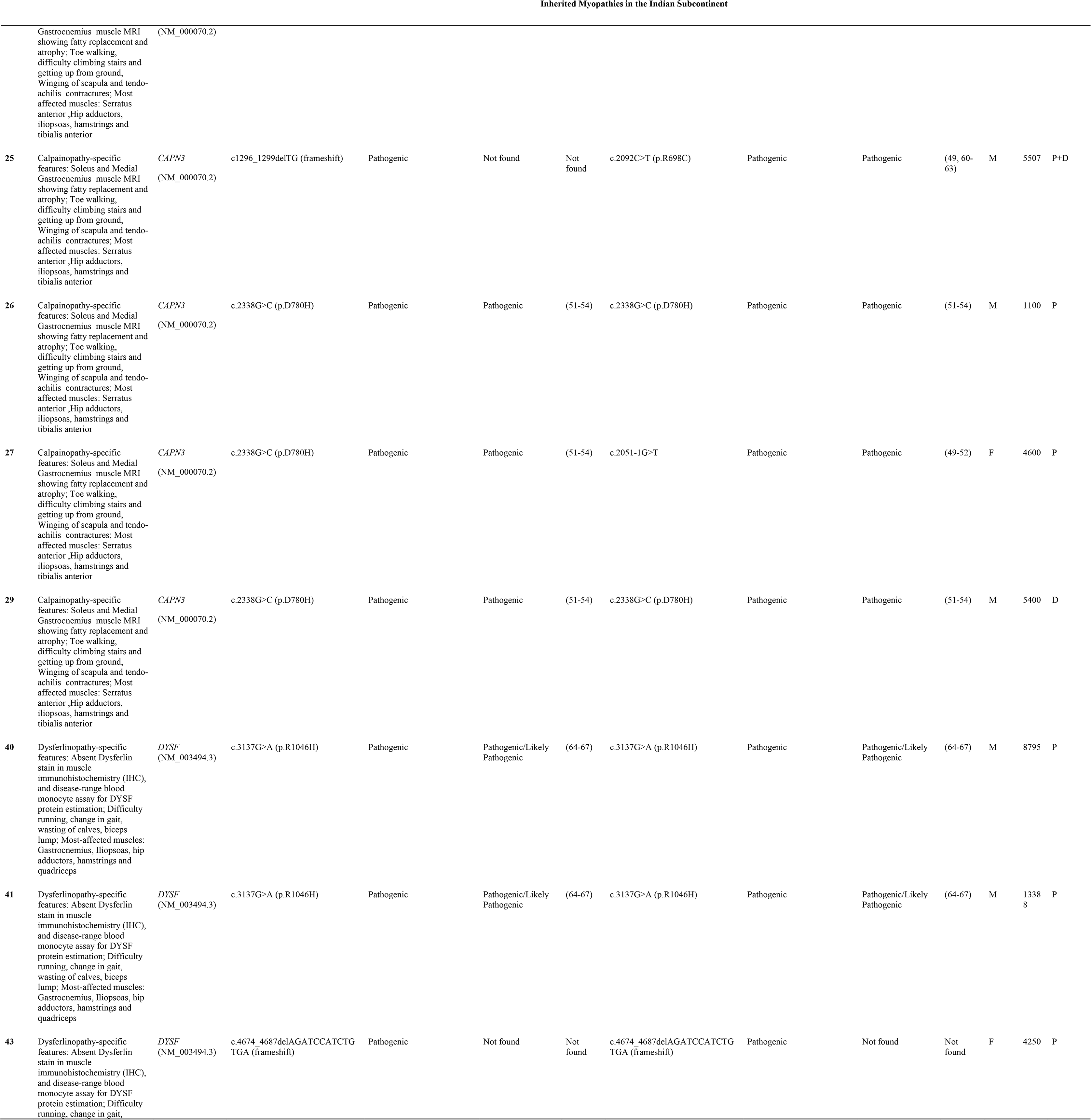
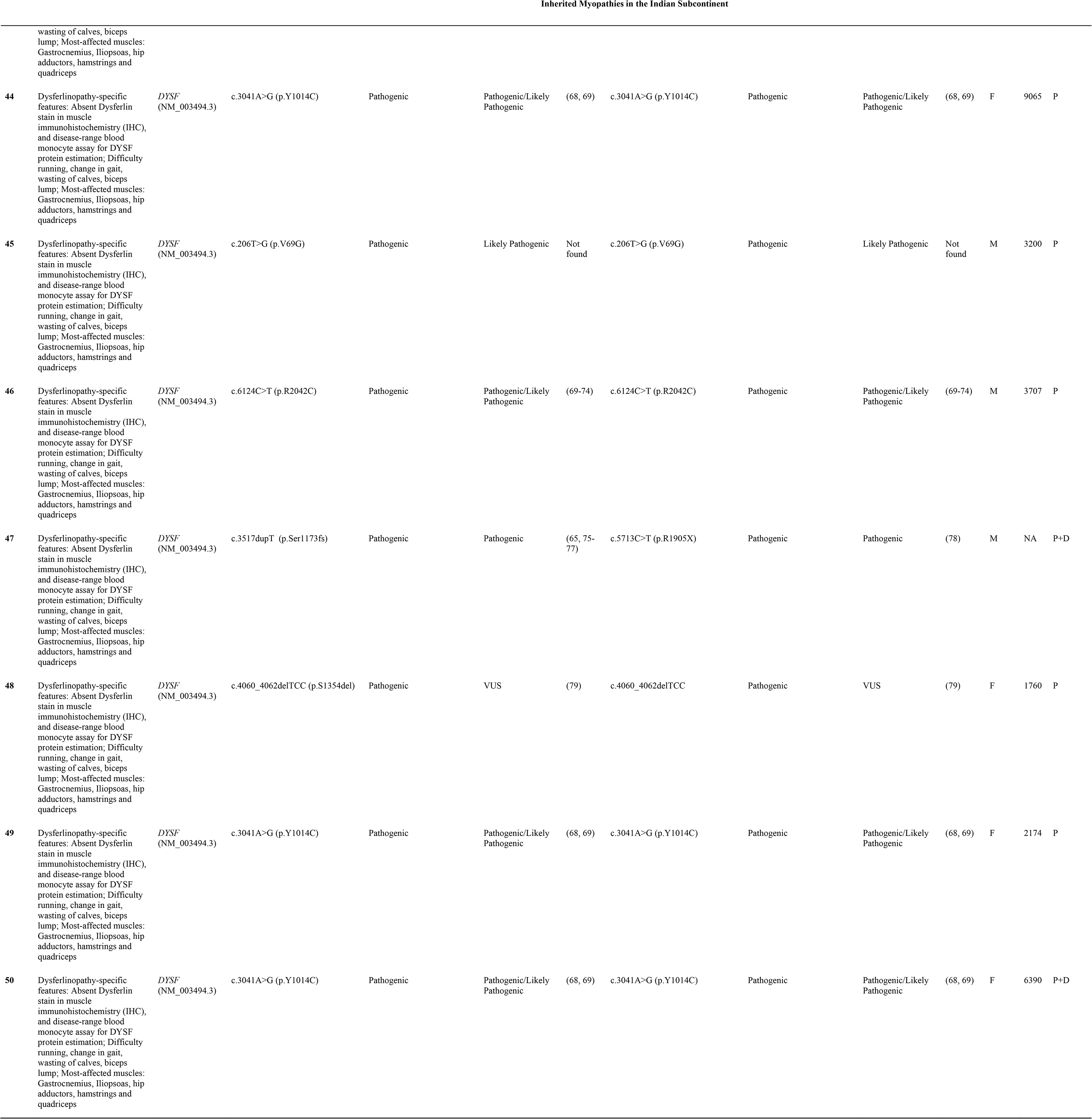
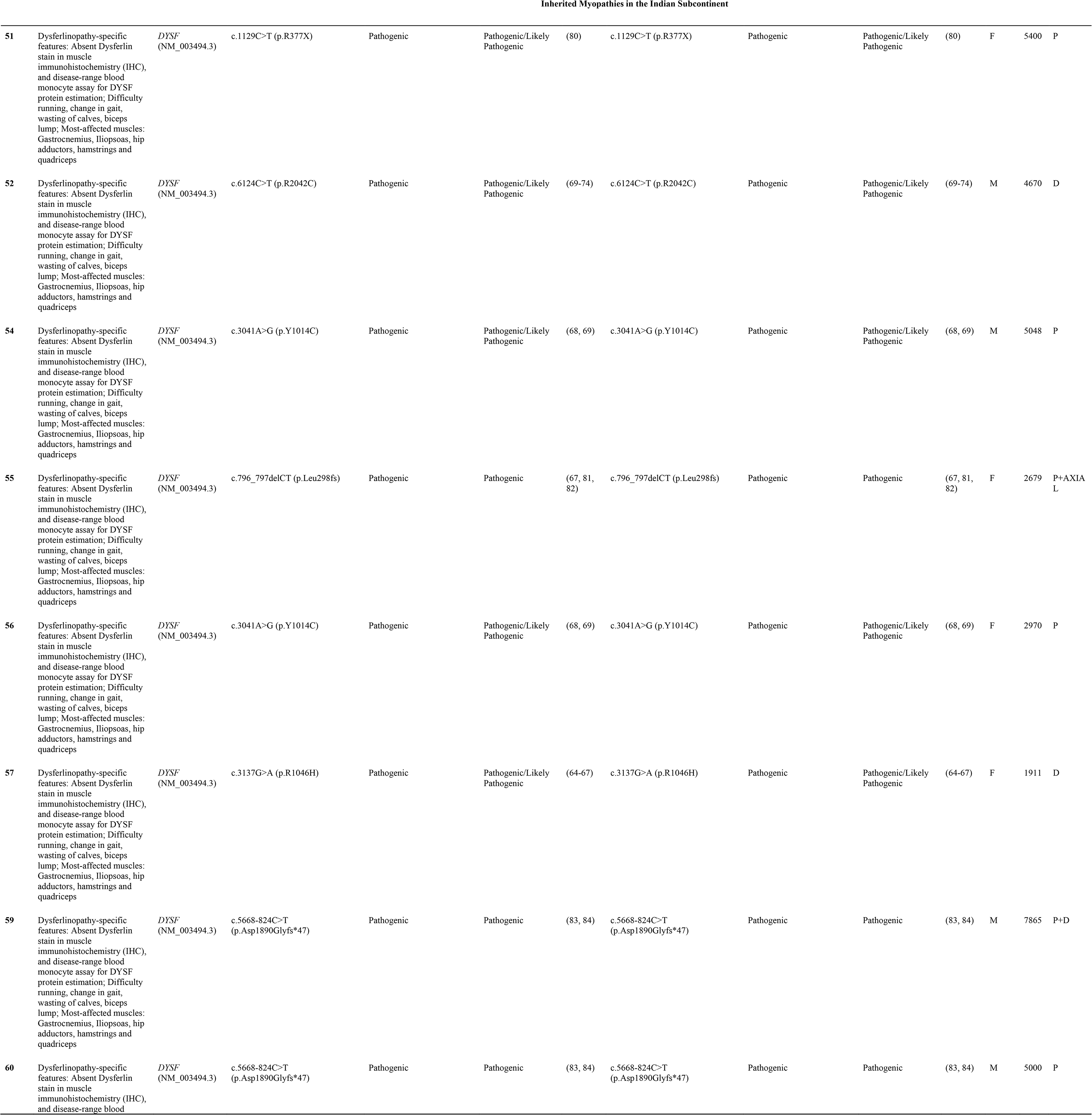
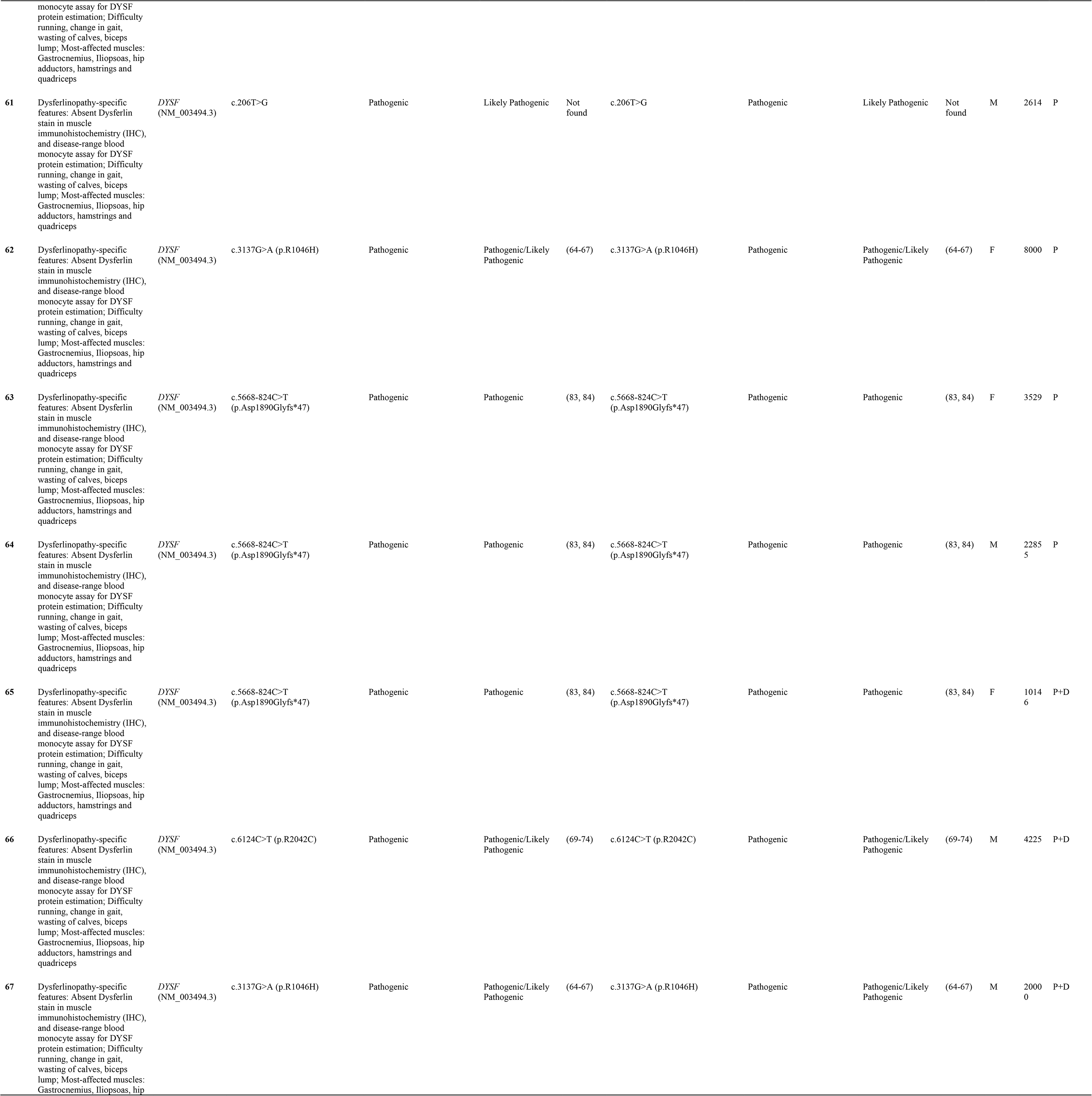
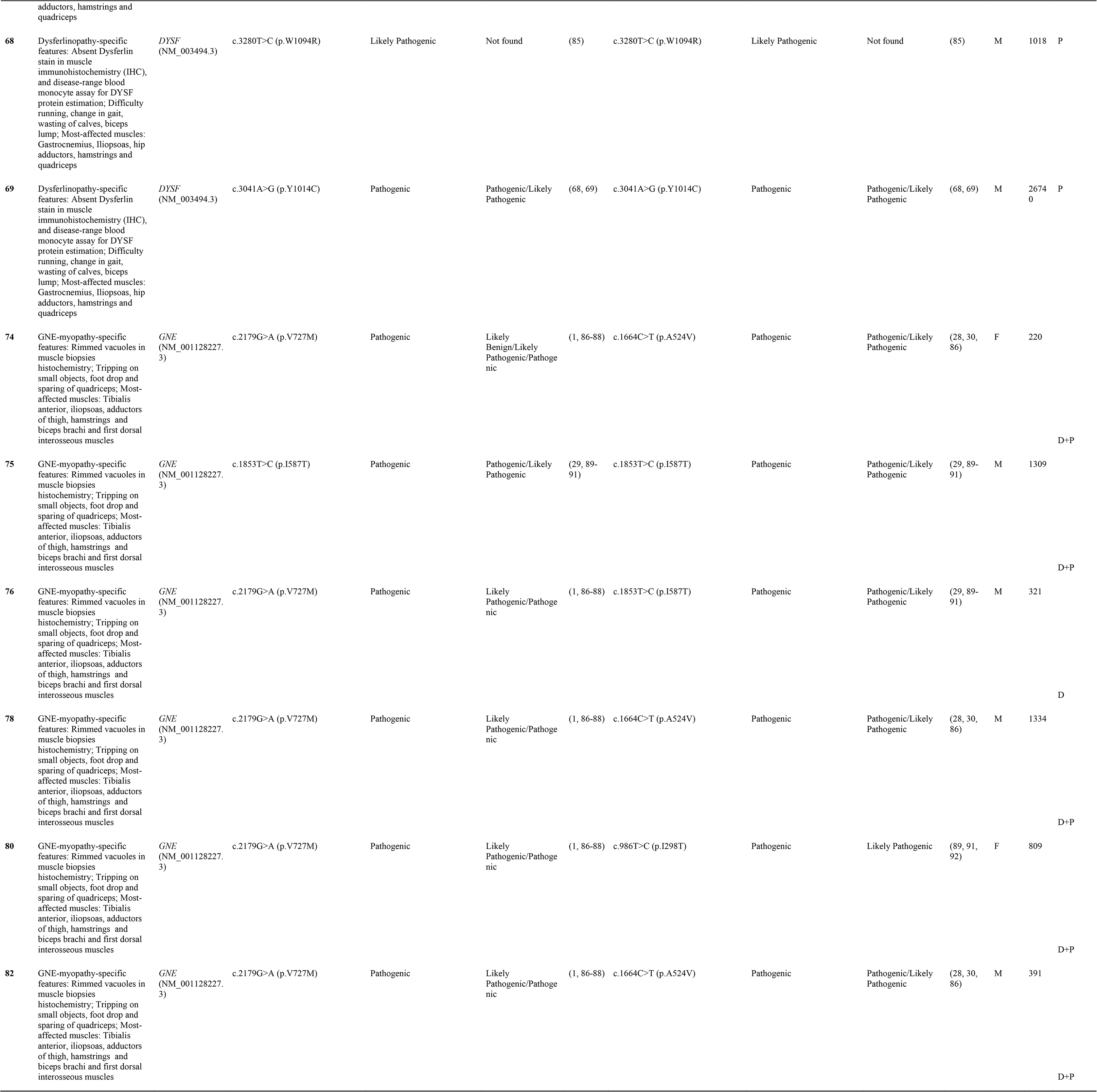
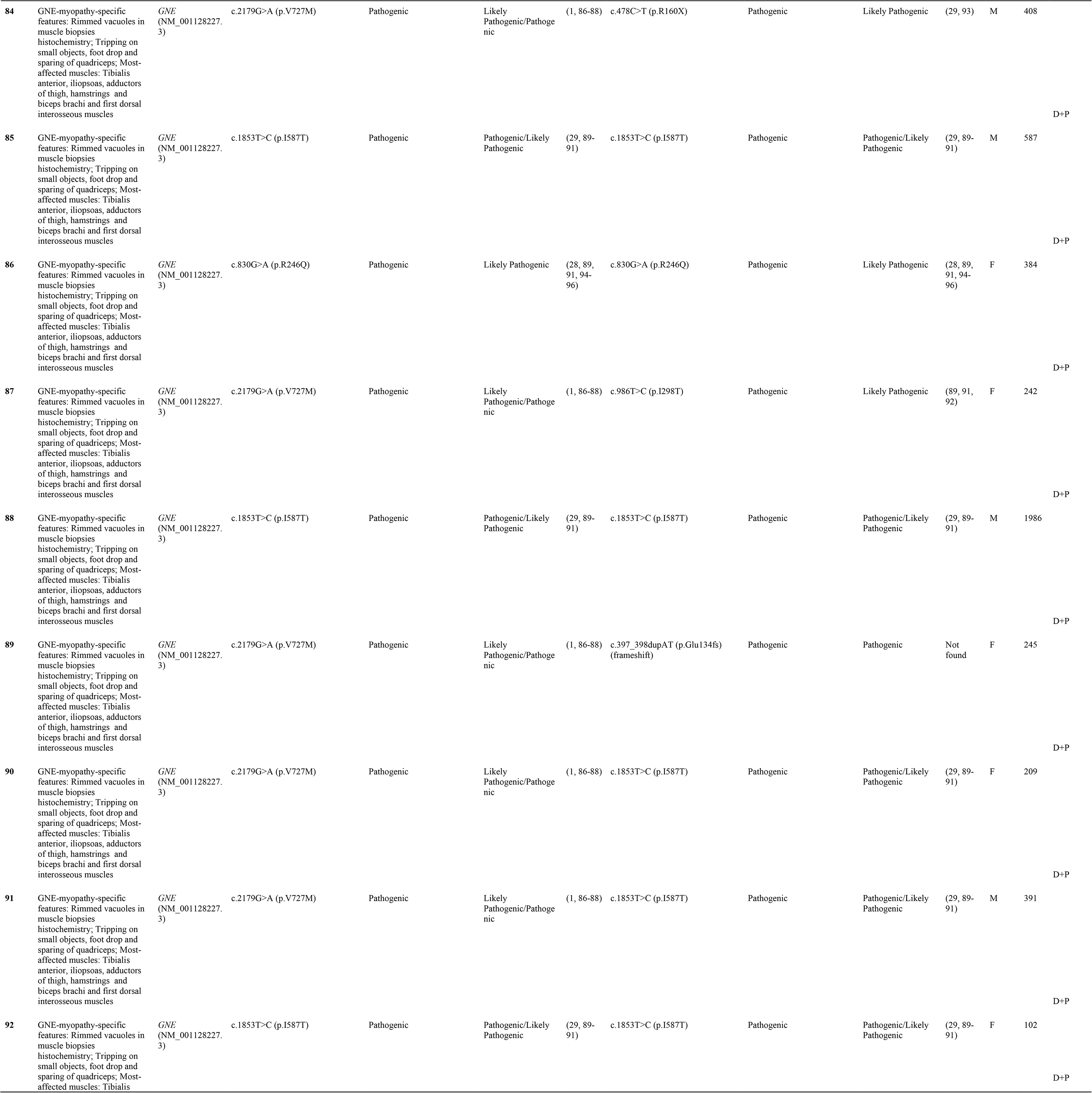
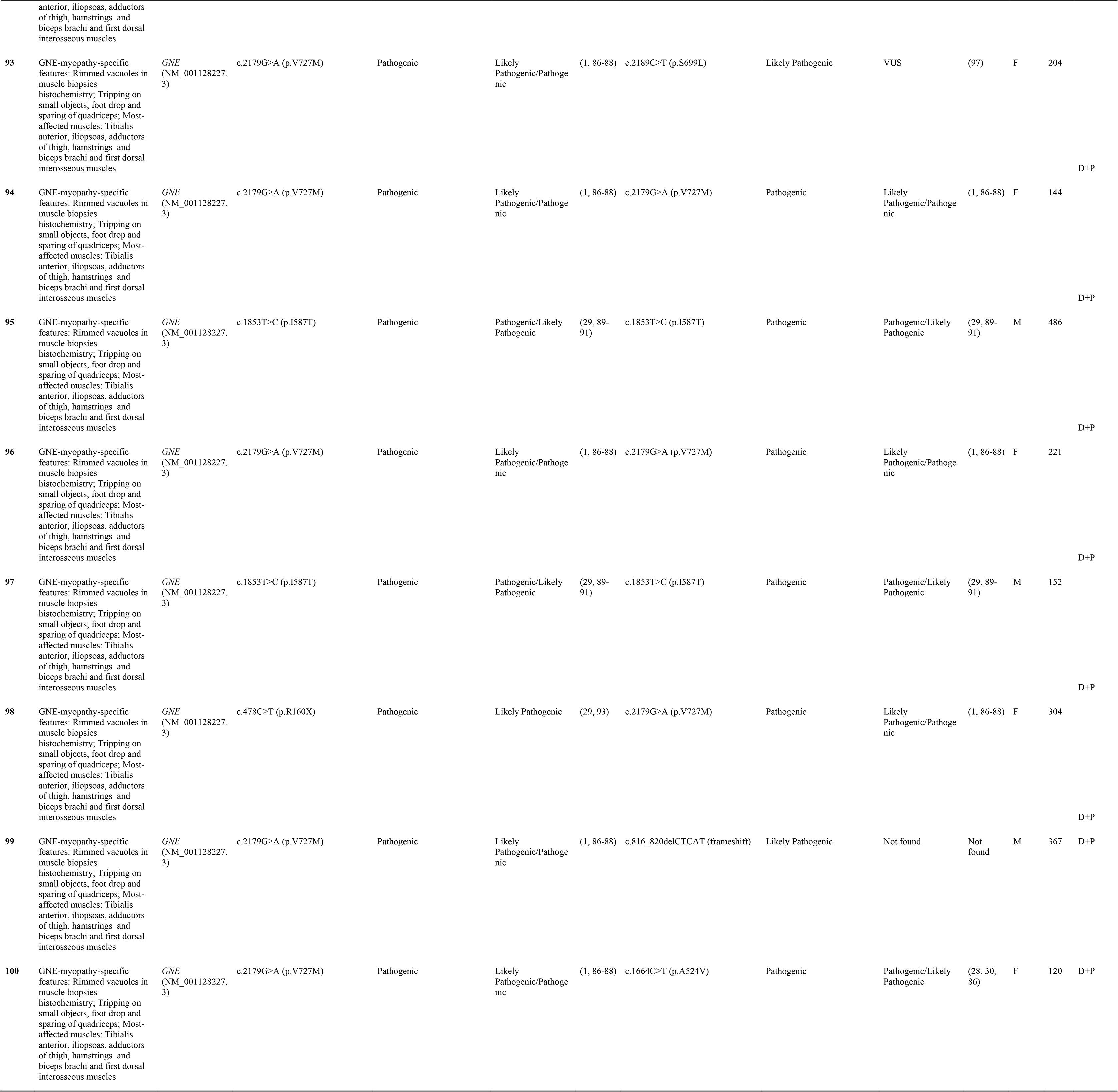
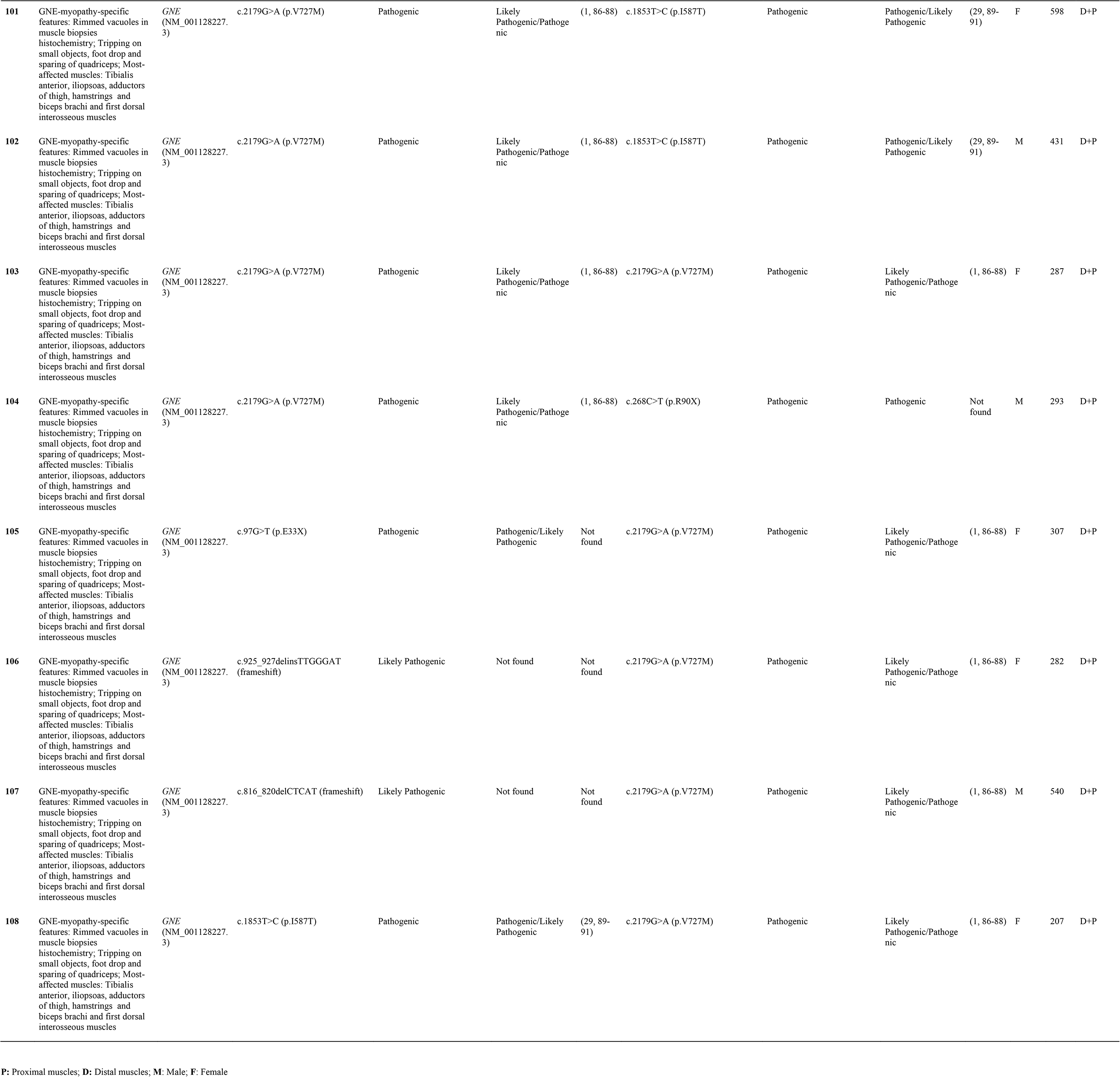
Molecular and Clinical aspects of *CAPN3, DYSF* and *GNE* patients.

**Figure 2.**
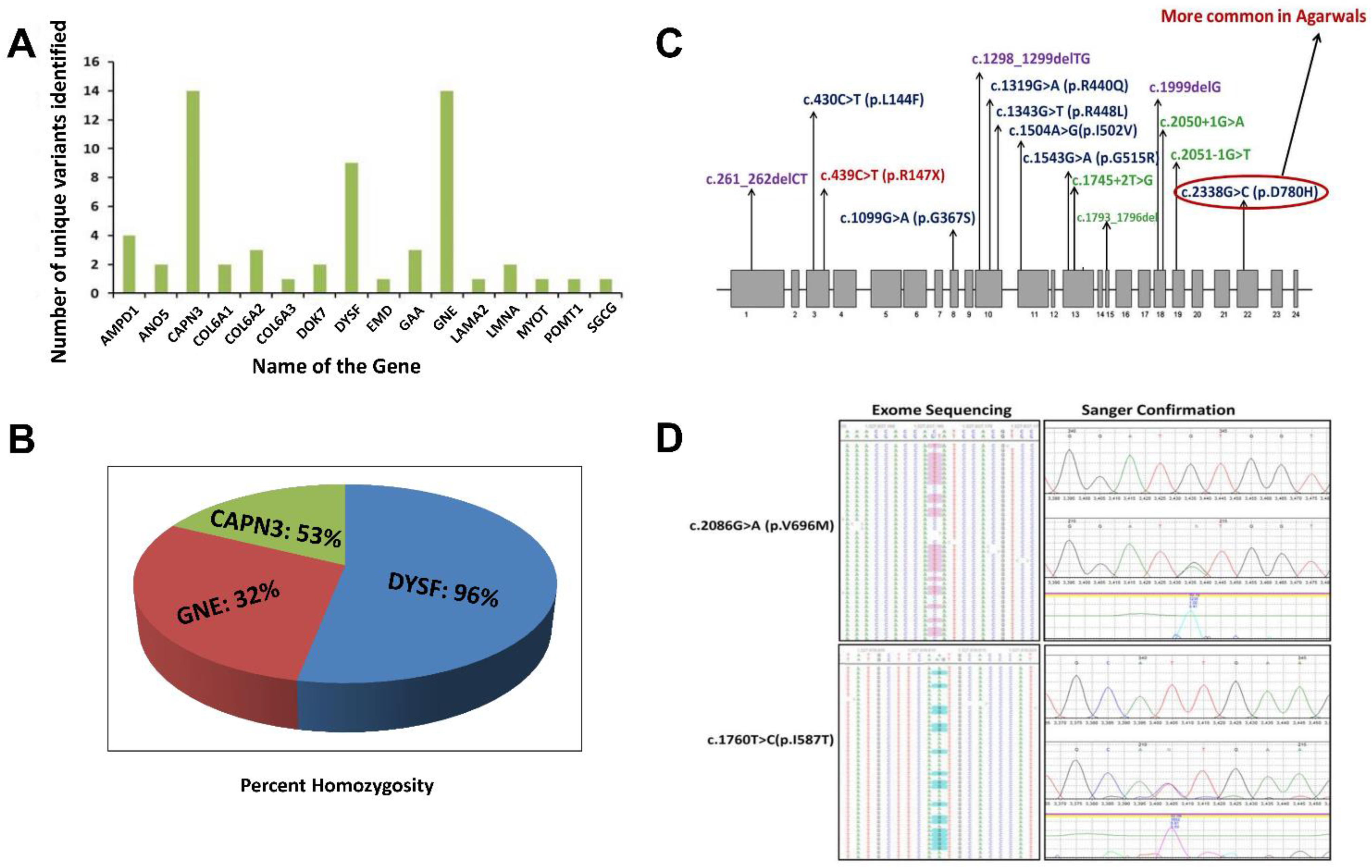
Unique pathogenic variants and percent homozygosity identified in major myopathy genes. A) Number of unique pathogenic variants identified in major myopathy genes. B) The percentage of homozygosity observed in the major myopathy subtypes. Among the 3 major myopathy subtypes, 96% of the limb-girdle muscular dystrophy R2 (LGMDR2) patients caused by variants in *DYSF* gene were identified with homozygous variants. C) Mutation spectrum of *CAPN3* gene: *CAPN3* variants identified in the current study are distributed throughout the gene. There is no specific mutation hot spot region but 3 variants are located in exon 10 indicating it as more unstable region prone to genetic variation. D) Two common variants in *GNE* gene: The two common variants in *GNE* gene were detected in exome sequencing with high confidence and also further confirmed by Sanger sequencing. Left side panel indicates the variants confirmation by exome sequencing. Right side panel indicates the variants confirmation by Sanger sequencing.

*CAPN3* and *DYSF* are the next major contributing genes in Indian genetic myopathy patients. Calpainopathy *(CAPN3*, LGMDR1) represents the most frequent LGMD-subtype worldwide (6, 20, 21), but was the second most common myopathy found in our cohort. No specific mutational hot spot was identified and all 14 identified unique variants in 9 patients were distributed throughout the *CAPN3* gene (Figure 2C). Homozygosity was observed in 4 out of 9 *CAPN3* patients (45%) (Figure 2A,B).

The 9 unique variants identified in 9 *DYSF* patients were also distributed throughout the *DYSF* gene. Interestingly, we found that 96% (26/27) of the individuals with a dysferlinopathy molecular diagnosis were homozygous for the detected pathogenic variant (Figure 2B), explaining why autosomal recessive disorders like dysferlinopathy are the more common form of inherited myopathy found in India.

In addition, the two Pompe genetically diagnosed Indian patients did not have either any homozygous *GAA* variants or the leaky splice-variant c.-32-13T>G, that are more common in adult-onset Pompe in many other populations.

### Clinical characteristics of genetically-diagnosed patients

Pictorial examples (Figure 3) and details of clinical features of common subtypes (Tables 1-2) and uncommon subtypes (Tables 3-4) of myopathies identified in our Indian cohort are provided. Cardiac involvement was observed in a total of 6 patients: one each for *AMPD1, GAA, DYSF, LMNA*, and two GNE-myopathies. Patient# 2 *(AMPD1)* and 38 *(LMNA:* Laminopathy or EDMD) showed atrioventricular conduction blocks, with AMPD1 patient showing 30% ejection fraction. Patient# 47 *(DYSF)* showed proximo-distal weakness and mild sub-clinical cardiomyopathy (ejection fraction 50%) resembling few reports of miyoshi myopathy (22, 23). Patient# 73 *(GAA:* Pompe disease) had dilated cardiomyopathy with 32% ejection fraction and also showed respiratory involvement with breathlessness on exertion. Patient # 89 and 90 (GNE-myopathies) showed both sub-clinical cardiomyopathy evidenced by mild reduction of ejection fraction on the 2D echocardiography. Among the 6 patients, only the patient # 73 (Pompe disease) showed breathlessness on exertion. Data on pulmonary function tests on any case were not available. Available muscle biopsy results indicated that 66% (31 out of 47) of the patients had dystrophic or myopathic changes with/without inflammation. A total of 4 patients were non-ambulatory and required wheel-chair assistance (Tables 1-4 and Table S1 in Supplementary Material).

**Figure 3.**
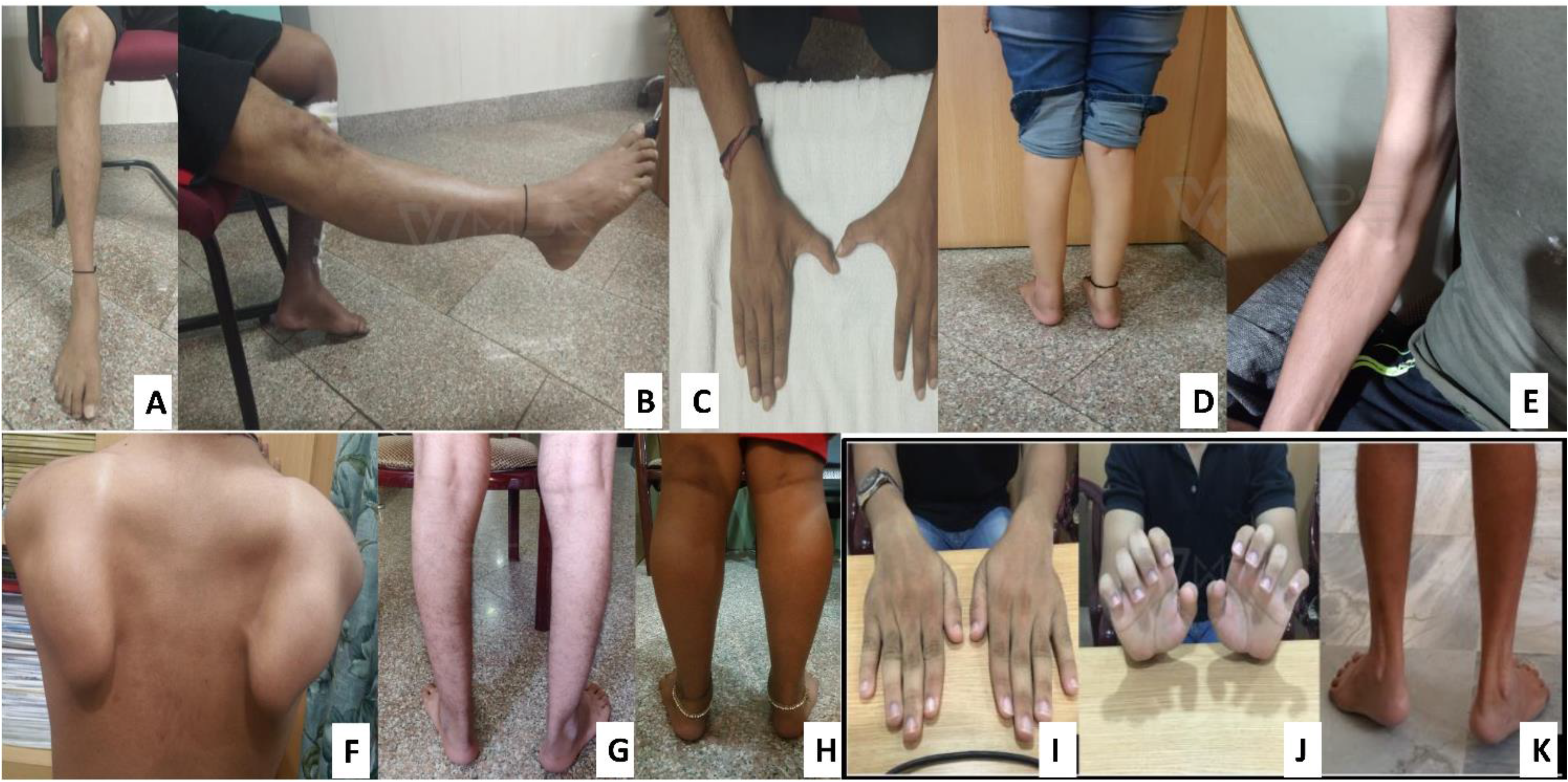
Classical features of common subtypes identified. GNE myopathy (A to C): Tibialis anterior (TA) muscle wasting, normal quadriceps and first dorsal interossei (FDI) muscle wasting identified respectively. Dysferlinopathy (D, E): Gastrocnemius wasting and biceps lump. Calpainopathy (F, G): Scapular winging and ankle contracture. Sarcoglycanopathy (H): Calf hypertrophy. Collagenopathy (I to K): Long finger flexor and ankle contractures.

**Table 2:** Clinical features of common subtypes.

| Parameters | Dysferlinopathy (LGMDR2) | Calpainopathy (LGMDR1) | GNE-myopathy |
| --- | --- | --- | --- |
| Number of patients | 27 | 19 | 31 |
| Mean age of onset(Range) | 22years (13-33) | 21 years (6 to 48 ) | 28 years (16-60) |
| Sex ratio (M:F) | 1.25:1 | 1.1:1 | 1.2:1 |
| Duration of illness(Range) | 1-17 years | 1-27 years | 3Months -30years |
| Commonest Presenting symptom | Difficulty running, change in gait | Toe walking, difficulty climbing stairs and getting up from ground | Tripping on small objects |
| Important Clinical markers | Wasting of calves (74%), Biceps lump ( 45%) | Winging of scapula (83%)and tendo-achilis contractures (71%) | Foot drop and sparing of quadriceps (100%). |
| Muscles most affected | Gastrocnemius, Iliopsoas, hip adductors, hamstrings and quadriceps | Serratus anterior ,Hip adductors, iliopsoas, hamstrings and tibialis anterior | Tibialis anterior, iliopsoas,adductors of thigh, hamstrings and biceps brachi and first dorsal interosseous muscles |
| Uncommon or Atypical features | Scapular winging and facial weakness (10%) | Diamond on quadriceps , biceps lump and facial weakness. | Scapular winging (6%) |
| Asymmetry at onset | 3% | - | 37% |
| CPK levels(Range) | 1018-26,740 IU/L | 690 to 7000 IU/L | 102-1986 IU/L. |

**Table 3.**
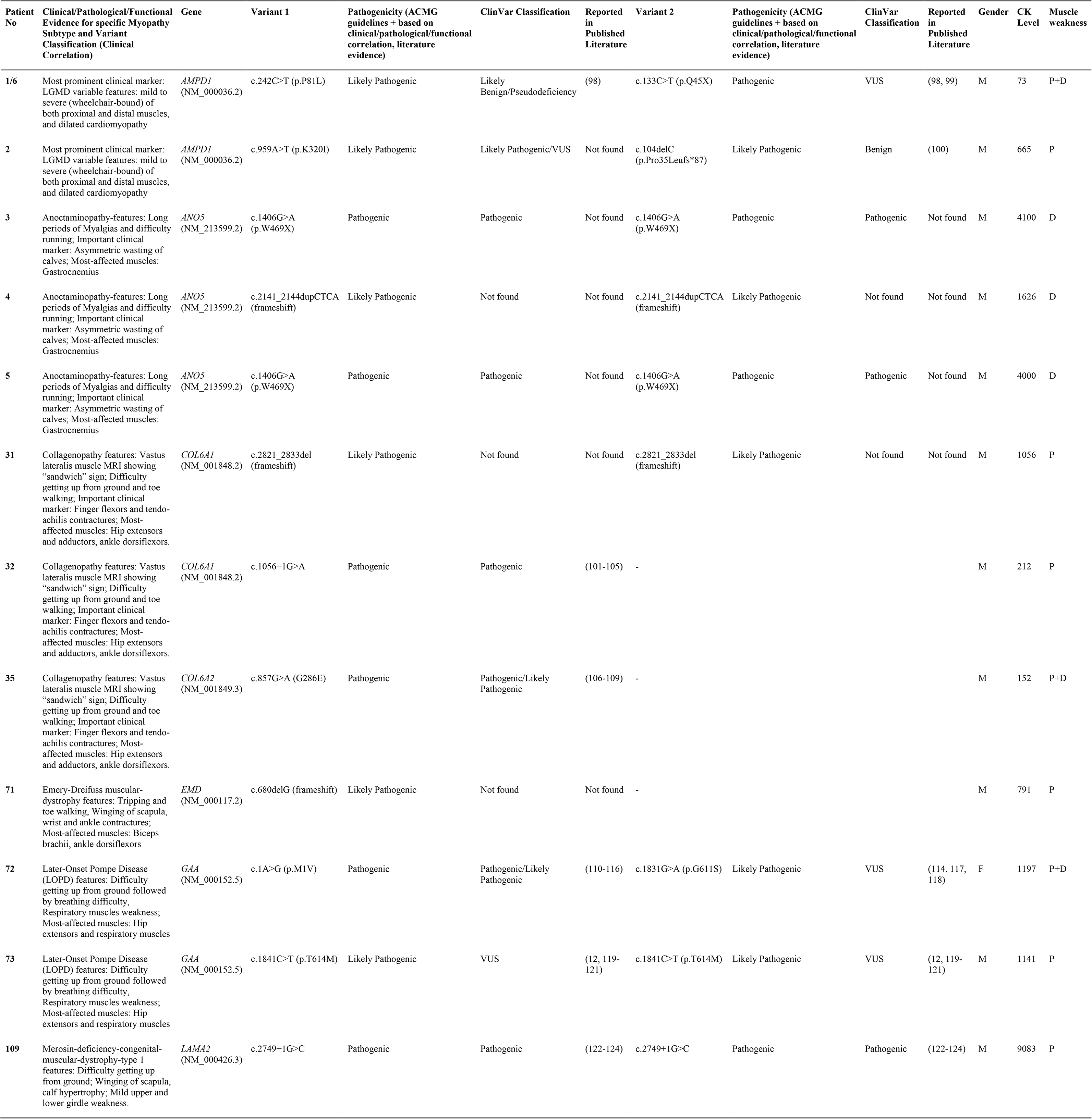
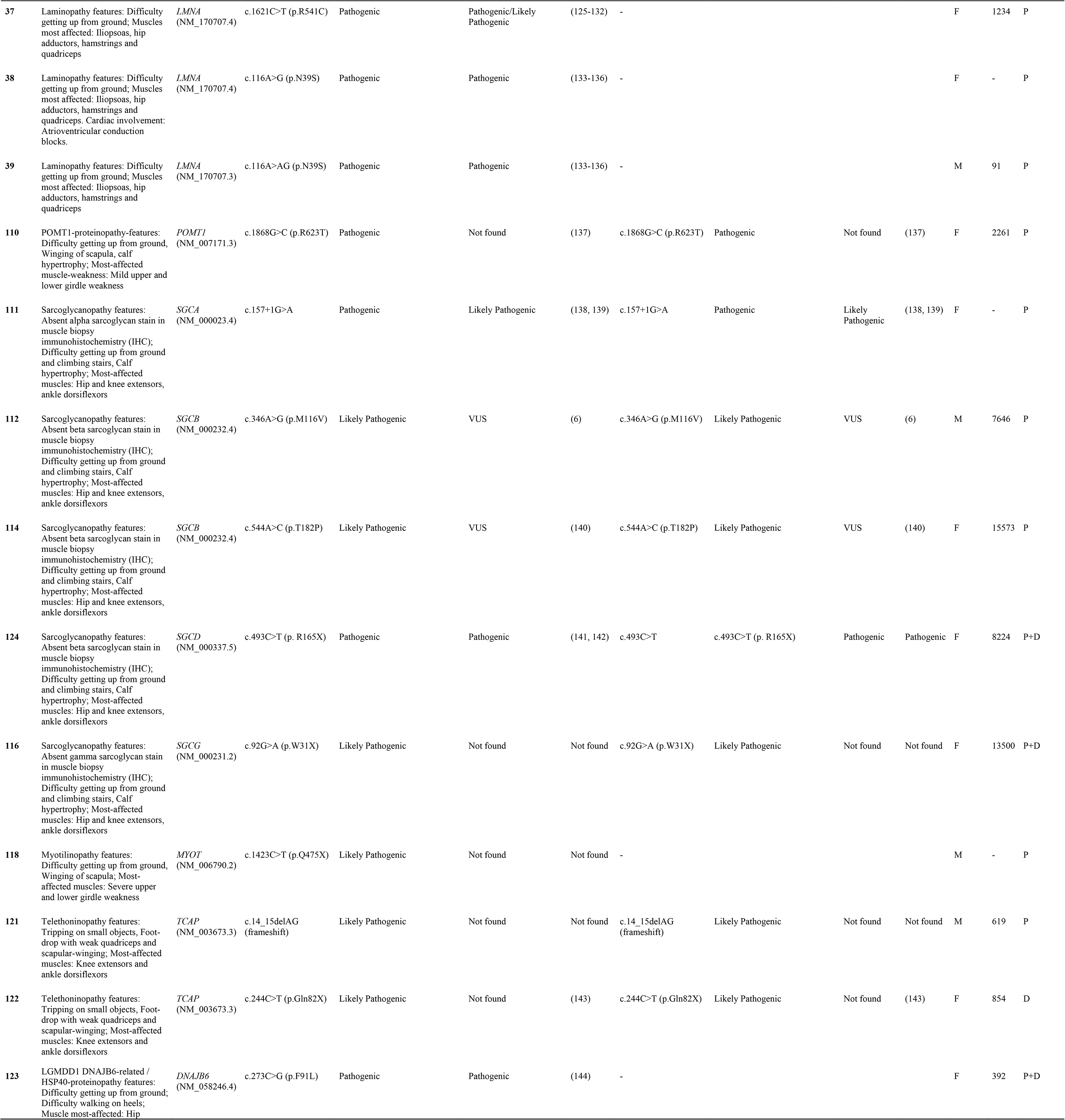
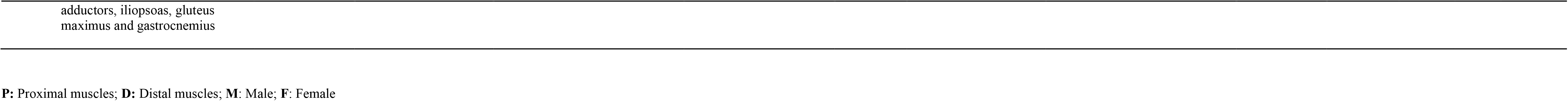
Uncommon subtypes identified in Indian patients.

**Table 4:**
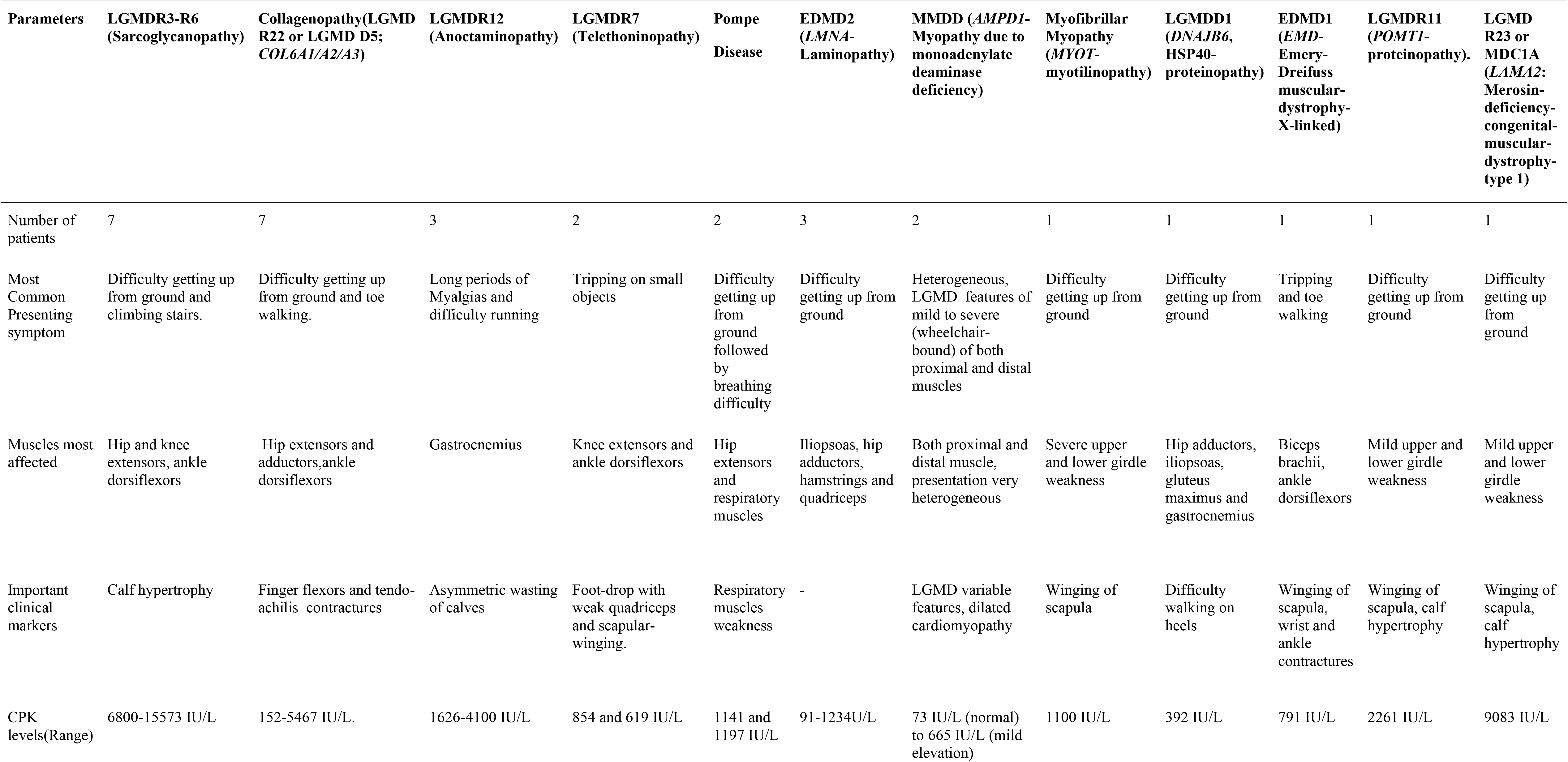
Clinical features of uncommon subtypes.

### Common Subtypes

GNE myopathy, dysferlinopathy and calpainopathy were identified as the three most common subtypes of inherited myopathies. Normal range of creatine kinase (CK) in this study was 30-160 IU/L. The details of the pattern of clinical weakness and uncommon clinical features of these three groups are described below.

### GNE-myopathy

The onset-age varied (16-60 years) in 17 females and 14 males. Total duration of illness was between 3 months and 30 years with lower limb onset in all except one with upper limb involvement simultaneously. Asymmetry at onset was seen in 13 patients. Creatine kinase (CK) levels ranged between 102-1986 IU/L. Two patients had loss of ambulation with long duration of illness (17 and 13 years).

#### Pattern of weakness

Most patients had conventional presentation with weakness affecting the tibialis-anterior (TA) maximally along with sparing of the quadriceps. With advancing duration of the disease, patients also showed a degree of weakness in the iliopsoas, adductors of thigh, hamstrings in lower-limbs and biceps and first dorsal interosseous muscles in the upper-limbs (denoted as D+P in Table 2).

#### Rare clinical features

Two patients had severe affection of proximal-muscles of the upper-limbs with winging, later into the illness (Table 2).

### Dysferlinopathy

Onset-age was 13-33 years in 15 males and 12 females. Duration of illness ranged from 1 to 17 years. Weakness began in the lower limbs of all patients: proximo-distal in 7 patients, only proximal in 21 patients, and only distal in the remaining 3 patients. Biceps lump was seen in 14 patients. CK levels highly varied and were between 1,018-26,740 IU/L.

#### Pattern of weakness

Most patients had proximal-onset of lower-limbs-weakness with maximum affection of iliopsoas, hip adductors, hamstrings, quadriceps, and gastrocnemius muscles. These patients had difficulty in walking on toes, wasting of calves and high CK-levels (Table 2).

#### Rare clinical features

Scapular winging and facial weakness was seen in 3 patients, in advanced stages of the disease. Asymmetry of weakness was seen in only 1 patient. This unusual patient had one pathogenic variant in the *ANO5* gene along with a homozygous pathogenic variant in *DYSF*. Interestingly, this patient (patient #50 in Table 5) showed a phenotype common to both dysferlinopathy and anoctaminopathy (high CK, proximal and distal muscle weakness, asymmetrical weakness) and features unusual to both types (facial weakness). Presence of inflammation in muscle biopsy and absence of Dysferlin staining were the manifestations of dysferlinopathy at the histological level.

**Table 5:**
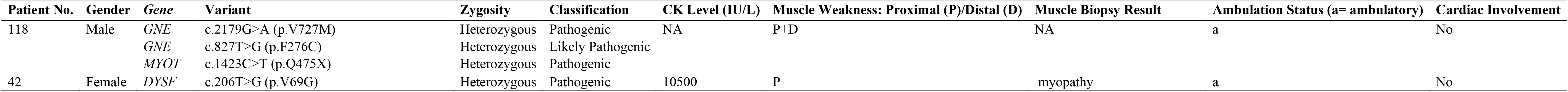
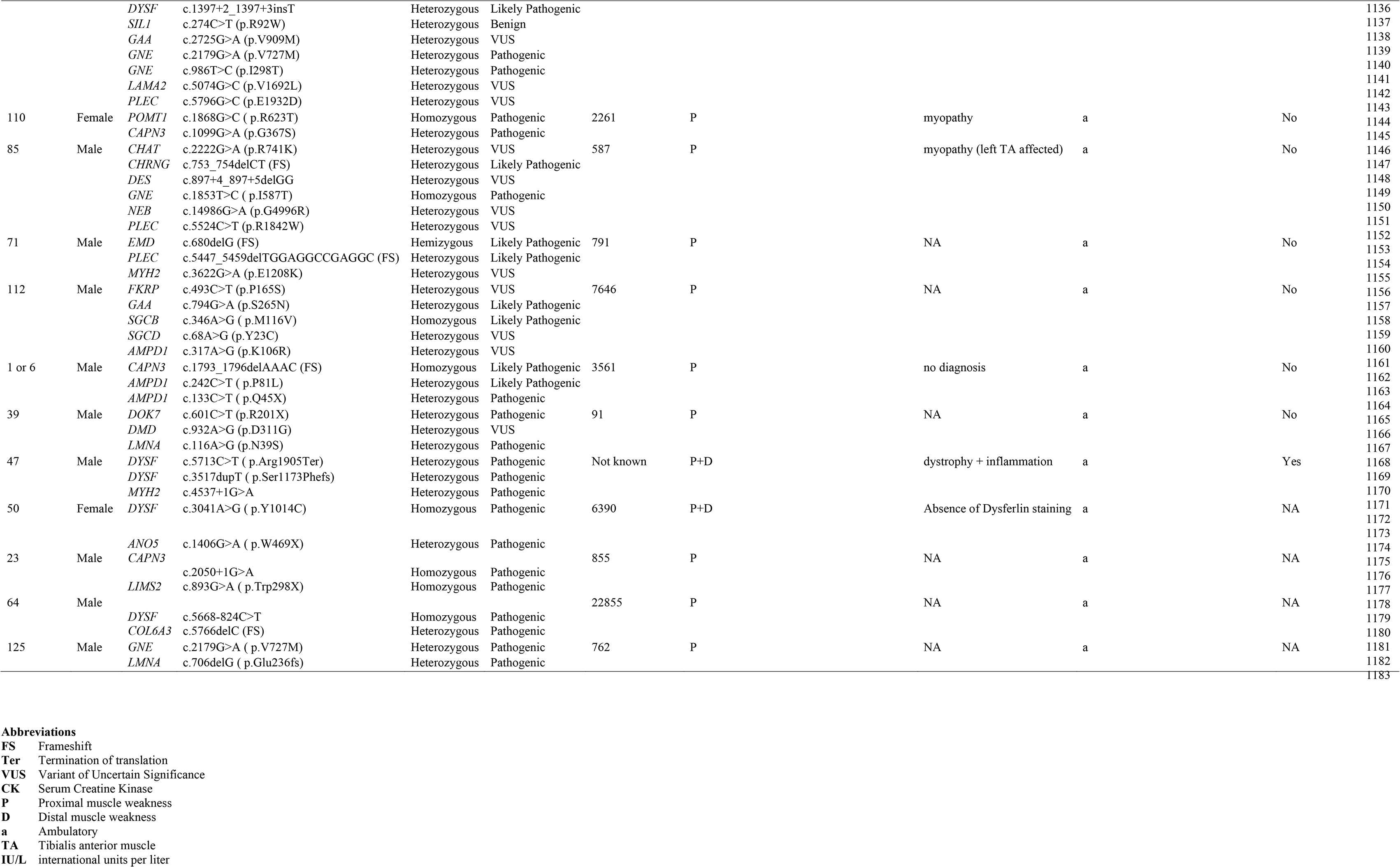
Clinical symptoms and genotype of patients with pathogenic variants in more than one gene.

### Calpainopathy

Onset-age was 6 to 48 years in 10 males and 9 females. Duration of illness ranged from 1 to 27 years. Lower limb onset of weakness was seen in 21-patients, upper limbs in 2 and upper and lower limbs were simultaneously affected in 1 patient. The weakness was proximal in 18, proximo-distal in 5 and distal in 1 patient. Winging was present in 15/19 and tendo-achilis contractures were seen in 12/19-patients. Loss of ambulation developed in one patient after 27years into the illness. CK levels ranged from 690 to 7000 IU/L.

#### Pattern of weakness

Most patients had the conventional proximal weakness-onset in the lower-limbs. Maximum weakness was seen in the hip adductors, iliopsoas, hamstrings, tibialis anterior (TA) and serratus anterior muscles. Tendoachillis contractures and toe-walking was common.

#### Rare clinical features

One patient had diamond on quadriceps and biceps lump which is commonly described in dysfelinopathy. One other patient had mild facial weakness.

### Clinical features of genetically-diagnosed uncommon subtypes in India

The clinical features of the rare and uncommon genetically diagnosed subtypes found in this study have been provided in Table 4. These individual groups were small consisting of few or a single patient each and were comprised of sarcoglycanopathies *(SGCA/SGCB/SGCD/SGCG)*, collagenopathies *(COL6A1/COL6A2/COL6A3)*, anoctaminopathy *(ANO5)*, telethoninopathy *(TCAP)*, Pompe disease (GAA), myoadenylate deaminase deficiency myopathy *(AMPD1)*, myotilinopathy *(MYOT)*, laminopathy *(LMNA)*, HSP 40 proteinopathy *(DNAJB6)*, Emery Dreifuss muscular dystrophy *(EMD)*, POMT1 proteinopathy *(POMT1)*, and merosin deficient congenital muscular dystrophy *(LAMA2)*. Interestingly, myoadenylate deaminase deficiency myopathy due to *AMPD1* pathogenic variants was only identified in 2 patients out of 207 Indian patient cohort (~1%) suggesting lower prevalence among individuals in the Indian subcontinent area and Asians in general. These 2 patients showed heterogeneous symptoms. One had mild limb girdle weakness with slightly elevated CK-levels (665 IU/L). The second patient was wheelchair bound but with normal CK levels (73 IU/L). Along with having compound heterozygous pathogenic *AMPD1* variants (c.242C>T and c.133C>T), the patient with the severe phenotype also harboured homozygous *CAPN3* pathogenic variant (c.1793_1796delAAAC; patient # 1 or 6), which could be associated with higher clinical severity. These *AMPD1* cases suggest more heterogeneity in the nature of muscle weakness and clinical overlap of *AMPD1*-associated myopathy with limb-girdle features in the Indian population.

We also identified 21 patients carrying VUSs in *CAPN3, DYSF, GNE, COL6A1, COL6A2, COL6A3, FLNC, SGCB, TRIM32* genes that has strong clinical correlation with the gene and corresponding myopathy subtype of interest but without further information reclassify them as likely pathogenic or pathogenic (Table 6). These cases are not included in the diagnosed patients in our cohort. Further segregation and functional studies are needed to confirm these VUSs’ pathogenicity.

**Table 6.** Molecular and Clinical aspects of patients (not included in diagnostic yield) with Variants of Uncertain Significance (VUSs) or Likely Pathogenic but with Clinical Correlation with Gene of interest.

| Patient No | Clinical/Pathological/Functional Evidence for specific Myopathy Subtype and Variant Classification (Clinical Correlation) | Gene | Variant 1 | Pathogenicity (ACMG guidelines + based on clinical/pathological/functional correlation, literature evidence)) | ClinVar Classification | Reported in Literature | Variant 2 | Pathogenicity (ACMG guidelines + based on clinical/pathological/functional correlation, literature evidence) | ClinVar Classification | Published Literature Evidence | Gender | CK Level | Muscle weakness |
| --- | --- | --- | --- | --- | --- | --- | --- | --- | --- | --- | --- | --- | --- |
| 9 | Calpainopathy-specific features: Soleus and Medial Gastrocnemius muscle MRI showing fatty replacement and atrophy; Toe walking, difficulty climbing stairs and getting up from ground, Winging of scapula and tendo-achilis contractures; Most affected muscles: Serratus anterior ,Hip adductors, iliopsoas, hamstrings and tibialis anterior | *CAPN3* (NM_000070.2) | c.1298_1299delTG (p.Val433fs) | Pathogenic | Pathogenic/Likely Pathogenic | (53) | c.1745+2T>G | VUS/Likely Pathogenic | Not found | Not found | F | 5440 | P+D |
| 10 | Calpainopathy-specific features: Soleus and Medial Gastrocnemius muscle MRI showing fatty replacement and atrophy; Toe walking, difficulty climbing stairs and getting up from ground, Winging of scapula and tendo-achilis contractures; Most affected muscles: Serratus anterior ,Hip adductors, iliopsoas, hamstrings and tibialis anterior | *CAPN3* (NM_000070.2) | c.430C>T (p.L144F) | VUS/Likely Pathogenic | Not found | Not found | c.430C>T (p.L144F) | VUS/Likely Pathogenic | Not found | Not found | F | NA | P |
| 11 | Calpainopathy-specific features: Soleus and Medial Gastrocnemius muscle MRI showing fatty replacement and atrophy; Toe walking, difficulty climbing stairs and getting up from ground, Winging of scapula and tendo-achilis contractures; Most affected muscles: Serratus anterior ,Hip adductors, iliopsoas, hamstrings and tibialis anterior | *CAPN3* (NM_000070.2) | c.439C>T (p.R147X) | Pathogenic | Pathogenic | Not found | c.1543G>A (p.G515R) | VUS/Likely Pathogenic | VUS | Not found | M | 1786 | P |

| Inherited Myopathies in the Indian Subcontinent |  |  |  |  |  |  |  |  |  |  |  |  |  |
| --- | --- | --- | --- | --- | --- | --- | --- | --- | --- | --- | --- | --- | --- |
| 17 | Calpainopathy-specific features: Soleus and Medial Gastrocnemius muscle MRI showing fatty replacement and atrophy; Toe walking, difficulty climbing stairs and getting up from ground, Winging of scapula and tendo-achilis contractures; Most affected muscles: Serratus anterior ,Hip adductors, iliopsoas, hamstrings and tibialis anterior | *CAPN3*<br>(NM_000070.2) | c.380-2A>G | VUS/Likely Pathogenic | Not found | Not found | c.380-2A>G | VUS/Likely Pathogenic | Not found | Not found | M | 4686 | P |
| 28 | Calpainopathy-specific features: Soleus and Medial Gastrocnemius muscle MRI showing fatty replacement and atrophy; Toe walking, difficulty climbing stairs and getting up from ground, Winging of scapula and tendo-achilis contractures; Most affected muscles: Serratus anterior ,Hip adductors, iliopsoas, hamstrings and tibialis anterior | *CAPN3*<br>(NM_000070.2) | c.1347C>A | VUS/Likely Pathogenic | Not found | Not found | c.1347C>A | VUS/Likely Pathogenic | Not found | Not found | F | 4226 | P |
| 42 | Dysferlinopathy-specific features: Absent Dysferlin stain in muscle immunohistochemistry (IHC), and disease-range blood monocyte assay for DYSF protein estimation; Difficulty running, change in gait, wasting of calves, biceps lump; Most-affected muscles: Gastrocnemius, Iliopsoas, hip adductors, hamstrings and quadriceps | *DYSF*<br>(NM_003494.3) | c.206T>G (p.V69G) | Pathogenic | Likely Pathogenic/VUS | Not found | c.1397+2_1397+3insT | VUS/Likely Pathogenic | VUS | Not found | F | 10500 | P |
| 53 | Dysferlinopathy-specific features: Absent Dysferlin stain in muscle immunohistochemistry (IHC), and disease-range blood monocyte assay for DYSF protein estimation; Difficulty running, change in gait, wasting of calves, biceps lump; Most-affected muscles: Gastrocnemius, Iliopsoas, hip adductors, hamstrings and quadriceps | *DYSF*<br>(NM_003494.3) | c.4168-1G>A | VUS/Likely Pathogenic | Not found | Not found | c.4168-1G>A | VUS/Likely Pathogenic | Not found | Not found | M | 17503 | D |
| 58 | Dysferlinopathy-specific features: Absent Dysferlin stain in muscle immunohistochemistry (IHC), and disease-range blood monocyte assay for DYSF protein estimation; Difficulty running, change in gait, wasting of calves, biceps lump; Most-affected muscles: Gastrocnemius, Iliopsoas, hip adductors, hamstrings and quadriceps | *DYSF*<br>(NM_003494.3) | c.3067A>T | VUS/Likely Pathogenic | Not found | Not found | c.4996delC (frameshift) | Likely Pathogenic | Not found | Not found | F | 6476 | P |
| 70 | Dysferlinopathy-specific features: Absent Dysferlin stain in muscle immunohistochemistry (IHC), and disease-range blood monocyte assay for DYSF protein estimation; Difficulty running, change in gait, wasting of calves, biceps lump; Most-affected muscles: Gastrocnemius, Iliopsoas, hip adductors, hamstrings and quadriceps | *DYSF*<br>(NM_003494.3) | c.2995T>C (W1017R) | VUS/Likely Pathogenic | VUS | Not found | c.2995T>C (W1017R) | VUS/Likely Pathogenic | VUS | Not found | M | 13502 | P+D |
| 77 | GNE-myopathy-specific features: Rimmed vacuoles in muscle biopsies histochemistry; Tripping on small objects, foot drop and sparing of quadriceps; Most-affected muscles: Tibialis anterior, iliopsoas, adductors of thigh, hamstrings and biceps brachi and first dorsal interosseous muscles | *GNE*<br>(NM_001128227.3) | c.2179G>A (p.V727M) | Pathogenic | Likely Pathogenic/Pathogenic | (1, 86-88) | c.1822C>A (p.P577T) | VUS/Likely Pathogenic | VUS | Not found | M | 799 | D+P |
| 79 | GNE-myopathy-specific features: Rimmed vacuoles in muscle biopsies histochemistry; Tripping on small objects, foot drop and sparing of quadriceps; Most-affected muscles: Tibialis anterior, iliopsoas, adductors of thigh, hamstrings and biceps brachi and first dorsal interosseous muscles | *GNE*<br>(NM_001128227.3) | c.2179G>A (p.V727M) | Pathogenic | Likely Pathogenic/Pathogenic | (1, 86-88) | c.95T>C (p.M32T) | VUS/Likely Pathogenic | VUS | Not found | M | 1880 | D+P |
| 81 | GNE-myopathy-specific features: Rimmed vacuoles in muscle biopsies histochemistry; Tripping on small objects, foot drop and sparing of quadriceps; Most-affected muscles: Tibialis anterior, iliopsoas, adductors of thigh, hamstrings and biceps brachi and first dorsal interosseous muscles | *GNE*<br>(NM_001128227.3) | c.2179G>A (p.V727M) | Pathogenic | Likely Pathogenic/Pathogenic | (1, 86-88) | c.1822C>A (p.P577T) | VUS/Likely Pathogenic | VUS | Not found | F | 210 | D+P |
| 83 | GNE-myopathy-specific features: Rimmed vacuoles in muscle biopsies histochemistry; Tripping on small objects, foot drop and sparing of quadriceps; Most-affected muscles: Tibialis anterior, iliopsoas, adductors of thigh, hamstrings and biceps brachi and first dorsal interosseous muscles | *GNE*<br>(NM_001128227.3) | c.124C>T (p.R11W) | Pathogenic | Likely Pathogenic | (145-148) | c.827T>G (p.F276C) | VUS/Likely Pathogenic | Not found | Not found | M | 1415 | D+P |
| 30 | Collagenopathy features: Vastus lateralis muscle MRI showing “sandwich” sign; Difficulty getting up from ground and toe walking; Important clinical marker: Finger flexors and tendo-achilis contractures; Most-affected muscles: | *COL6A1*<br>(NM_001848.2) | c.1848_1850delCCT (p.L617del) | VUS/Likely Pathogenic | VUS | Not found | c.1848_1850delCCT (p.L617del) | VUS/Likely Pathogenic | VUS | Not found | M | 455 | P+D |
|  | Hip extensors and adductors, ankle dorsiflexors. |  |  |  |  |  |  |  |  |  |  |  |  |
| 33 | Collagenopathy features: Vastus lateralis muscle MRI showing “sandwich” sign; Difficulty getting up from ground and toe walking; Important clinical marker: Finger flexors and tendo-achilis contractures; Most-affected muscles: Hip extensors and adductors, ankle dorsiflexors. | COL6A2 (NM_001849.3) | c.1591G>A (p.G531S) | VUS/Likely Pathogenic | VUS | Not found | c.1970-9G>A | Pathogenic | Pathogenic/Likely Pathogenic | (149-153) | M | 5467 | P |
| 34 | Collagenopathy features: Vastus lateralis muscle MRI showing “sandwich” sign; Difficulty getting up from ground and toe walking; Important clinical marker: Finger flexors and tendo-achilis contractures; Most-affected muscles: Hip extensors and adductors, ankle dorsiflexors. | COL6A2 (NM_001849.3) | c.1900G>A (p.E634K) | VUS/Likely Pathogenic | Not found | Not found | c.1970-2A>G | VUS/Likely Pathogenic | Not found | Not found | F | 215 | P |
| 36 | Collagenopathy features: Vastus lateralis muscle MRI showing “sandwich” sign; Difficulty getting up from ground and toe walking; Important clinical marker: Finger flexors and tendo-achilis contractures; Most-affected muscles: Hip extensors and adductors, ankle dorsiflexors. | COL6A3 (NM_004369.3) | c.8301_8303delCTT (p.Y2767_F2768del) | VUS/Likely Pathogenic | Not found | Not found | c.8301_8303delCTT (p.Y2767_F2768del) | VUS/Likely Pathogenic | Not found | Not found | F | 615 | P+D |
| 119 | Filaminopathy-features: Difficulty getting up from ground, Winging of scapula; Most-affected muscles: Mild Upper and lower girdle weakness | FLNC (NM_001458.4) | c.5278G>A (p.G1760S) | Likely Pathogenic | VUS | (10, 154) | c.6043G>C (p.V2015L) | VUS/Likely Pathogenic (South Asia MAF: 0.0061%) | Not found | Not found | M | 500 | P |
| 113 | Sarcoglycanopathy features: Absent beta sarcoglycan stain in muscle biopsy immunohistochemistry (IHC); Difficulty getting up from ground and climbing stairs, Calf hypertrophy; Most-affected muscles: Hip and knee extensors, ankle dorsiflexors | SGCB (NM_000232.4) | c.544A>C (p.T182P) | Likely Pathogenic | VUS | (140) | c.181A>G (p.K61E) | VUS/Likely Pathogenic (novel) | Not found | Not found | F | 6800 | P |
| 115 | Sarcoglycanopathy features: Absent beta sarcoglycan stain in muscle biopsy immunohistochemistry (IHC); Difficulty getting up from ground and climbing stairs, Calf hypertrophy; Most-affected muscles: Hip and knee extensors, ankle dorsiflexors | SGCB (NM_000232.4) | c.335A>T | VUS/Likely Pathogenic (novel) | Not found | Not found | c.335A>T | VUS/Likely Pathogenic (novel) | Not found | Not found | F | 7650 | P |
| 117 | TRIM32-proteinopathy features: Difficulty getting up from ground, Calf hypertrophy; Most-affected muscles: Mild upper and lower girdle weakness | TRIM32 (NM_001099679.2) | c.59G>T (p.C20F) | VUS/Likely Pathogenic | Not found | Not found | c.59G>T (p.C20F) | VUS/Likely Pathogenic | Not found | Not found | F | 1708 | P+D |

### Multigenic myopathies: a novel disease mechanism

Interestingly, 13 patients were identified with pathogenic variants in more than one myopathy genes (Table 5) all showing unusual presentations. We describe below such cases of multigenic unusual presentations identified in this study.

Patient# 118 with one pathogenic variant each in the *GNE* (OMIM# 603824) and myotilin *(MYOT;* OMIM#604103) genes. His phenotype largely resembled GNE-myopathy with foot-drop and quadriceps-sparing. However, disease-onset was both proximal and distal which also could resemble both subtypes. By the time foot-drop started, difficulty in rising from ground and low chairs had already developed. In this patient, both the genetic abnormalities could be contributing to the proximo-distal presentation resembling both autosomal-dominant myotilinopathy (24) as well as autosomal-recessive GNE-myopathy that has been reported to show early onset of proximal weakness mimicking LGMD especially with hamstring weakness (25). Muscle biopsy staining showed rimmed vacuoles, a common feature of both GNE-myopathy and *MYOT-related* myofibrillar myopathy. Myotilin immunostaining could have helped further but was not available.

Patient# 42 had pathogenic variants in the *GNE* gene and additionally harbored one likely pathogenic variant in the dysferlin *(DYSF)* gene. The phenotype was strongly suggestive of a GNE-myopathy, but had onset of proximal muscle weakness and presented a high CK level (1050 IU/L) which are a very well-known presentation of dysferlinopathy suggesting blended phenotype. GNE-myopathy patients are known to have proximal weakness only in later stages of illness. Muscle biopsy confirmed rimmed vacuoles but immunocytochemistry for dysferlin was not available, resembling GNE-myopathy more than dysferlinopathy.

Patient# 110 showed the most unusual symptoms: quadriceps-sparing and foot drop resembling GNE-myopathy, but with early and prominent development of scapular-winging and calf-hypertrophy. Interestingly, a heterozygous pathogenic variant in *CAPN3* (OMIM# 114240) and homozygous pathogenic variant in *POMT1* (OMIM# 607423) were present without any *GNE* variant. While the *CAPN3* variant can explain adductor, posterior thigh weakness and scapular winging, *POMT1* variants are known to be associated with hypertrophic calves (26) in limb-girdle muscular dystrophy (LGMDR11). Quadriceps-sparing and prominent foot-drop are unusual in the demonstrated genetic abnormalities.

Another patient (patient# 50) with a homozygous pathogenic variant in *DYSF* and a heterozygous pathogenic variant in *ANO5*, showed phenotypes of dysferlinopathy and anoctaminopathy that are representative of both subtypes (calf atrophy, CK of 6390 IU/L, combination of proximal and distal weakness, presence of inflammation in muscle-biopsy, asymmetry of muscle weakness, weakness in dorsiflexors of ankle joints, partial foot drop), and feature unusual to both (facial weakness).

We identified one patient clinically diagnosed with LGMDR2 but with an unexpected severe progression. This patient had pathogenic variants in both *DYSF* and *MYH2* genes (patient# 47). Heterozygous loss of *MYH2* coupled with complete loss of *DYSF* could be contributing to the rapid disease progression and severe phenotype of both proximal and distal muscle weakness along with cardiac-involvement observed in this individual.

Patient# 23 showed homozygous pathogenic variants in both *CAPN3* and *LIMS2* genes suffered from progressive weakness of hip and shoulder girdles. The phenotype consisted of scapular winging, lordosis, severe weakness of hip adductor muscles and mild tendo-achillis contractures, resembling calpainopathies. He also had mild calf hypertrophy. While transient or persistent calf hypertrophy has been documented in calpainopathies(27), it is also a feature of the *LIMS2*-patients. He did not have the triangular tongue or cardiomyopathy seen in *LIMS2*-patients.

Patient# 125 had one pathogenic variant each in both *LMNA* and *GNE* genes with limb-girdle pattern of weakness with calf hypertrophy and subclinical cardiac involvement in the form of atrioventricular conduction block. CK level was mildly raised (762 IU/L). All these features are in favor of autosomal-dominant laminopathy. Examination did not reveal any features suggestive of GNE-Myopathy but since *GNE* variants can be segregated either recessively or dominantly, longitudinal patient and family natural history is being followed through.

Similar multigenic combinations of pathogenic variants were detected for *GNE, ANO5, MYOT, CAPN3, LIMS2, COL6A3, POMT1, CHRNG, EMD, PLEC, GAA, SGCB, DMD, DOK7*, and *LMNA* genes in a total of 13 individuals (Table 5).

## DISCUSSION

Most of the inherited myopathies impose difficulties with physical activity, walking, poor quality of life and ultimately cause a heavy burden on both the affected individuals and their families. Identifying the correct diagnosis of these inherited myopathies can aid in disease management, treatment, and family planning. Clinical diagnosis is based on the distribution of predominant muscle weakness, inheritance mode and associated symptoms but is often highly elusive due to the overlap in clinical presentation. Therefore molecular diagnosis is necessary to confirm the identifying disease-causative gene. This study provides for the first time molecular diagnosis and clinical information on a fairly large cohort-size of genetic myopathies in patients of diverse ethnic backgrounds seen in the Indian subcontinent.

This cohort includes a large variety of inherited myopathies including GNE-myopathy, varieties of LGMDs, collagenopathies, metabolic myopathies and related others which have a significant phenotypic-overlap. *GNE, DYSF*, and *CAPN3* are the three major genetic contributors to these myopathies, in the Indian subcontinent. Even though we recruited patients from across the Indian subcontinent, as the evaluation was performed in a major single center hospital, there could be limitations to the interpretation of the prevalence. With upcoming or ongoing interventional or natural history study clinical trials on these myopathy subtypes (https://clinicaltrials.gov/ct2/results?recrs=ab&cond=GNE+Myopathy+&term=&cntry=&state=&city=&dist=,HYPERLINK "https://www.sarepta.com/science/gene-therapy-engine" https://www.sarepta.com/science/gene-therapy-engine), this study will enable cohorts from Indian subcontinent to be included in patient registries that, in turn, will enhance the clinical trials by including different populations with varied ethnicities as well as better monitoring of trial efficacies. Our diagnostic yield of 49% is considerably high with careful clinical correlation of the gene and rare variant identified. Another reason for a high diagnostic yield in an Indian subcontinent cohort is the high prevalence of consanguinity leading to greater homozygosity as found in all the three major contributing genes: *GNE, DYSF*, and *CAPN3* (Figure 2B). Moreover, our careful clinical pre-screening with inclusion criteria of genetic myopathies and excluding DMD, FSHD, DM1, DM2, mitochondrial myopathies and acquired myopathies enabled a higher diagnostic yield and further suggests the importance of phenotype correlation of clinical genetic testing results. The clinical features of the three major genetic myopathies followed those mentioned in the literature to a large extent such as GNE myopathy patients with weakness of the tibialis anterior muscles with sparing of quadriceps, dysferlinopathy patients having gastrocnemius weakness and proximal weakness, and calpainopathies having winging of scapulae and hip girdle weakness, mainly of adductor muscles. In addition, GNE-myopathy patients in our cohort also had proximal weakness, possibly representing advanced stages of the disease. The clinical features of the common and uncommon types are summarised in Tables 2 and 4, respectively.

Molecular findings related to GNE-myopathy patients identified in this study have been reported recently (28) with the most common pathogenic variant in our cohort being the c.1760T>C/c.1853T>C (p.I587T/p.I618T). This variant has recently been described with an ethnic founder effect (29, 30). Moreover, p.V727M is potentially a founder variant in Indian subcontinent since it was seen with high prevalence in our study and previous studies (31-33), and hence this variant is likely present in carriers at higher rates in the Indian subcontinent. This suggests the need for family tracing and carrier testing for inherited rare disorders such as GNE myopathy.

Calpainopathy (LGMDR1) was considered a strict autosomal-recessive LGMD-subtype for many years, but patients carrying specific single pathogenic deletion variants in the *CAPN3* gene are reported recently showing dominant disease segregation (6, 34) which was not identified in Indian patients in the current study.

Limb-girdle muscular dystrophy type 2B (LGMDR2) and Miyoshi myopathy (MM) caused by variants in the dysferlin gene, *DYSF* (35) are the two major clinical types of dysferlinopathy (36), characterized by proximal muscle-weakness, difficulty in running and climbing stairs, and increased fatigue (37). The higher (96%) homozygosity rate of pathogenic variant in *DYSF* gene explains why autosomal recessive disorders like dysferlinopathy are more common in Indian subcontinent. *DYSF* gene is much larger than some of the other genes such as *GNE* and *CAPN3*, the other two most prevalent myopathy forms in this study. Thus *DYSF* likely harbors more variants and therefore has a higher chance of homozygosity.

In this cohort many other more rare genetic myopathies were seen (Tables 3-4, Table S1 in Supplementary Material). Out of these, sarcoglycanopathies and collagenopathies were seen more frequently than other myopathies. The phenotypes of most myopathies were comparable to literature descriptions. There were some unusual clinical features and heterogeneity as well, such as that in *AMPD1* associated myopathy where ranges from mild symptoms to severe limb-girdle features progressing fast to wheel-chair assistance and dilated cardiomyopathy were observed. Only 2 *AMPDL*-associated myopathy cases were identified in our Indian subcontinent cohort suggesting lower prevalence in Asia similar to what was shown before in the Japanese population(38), compared to being one of the most common genetic myopathies among Caucasians (1 in 50-100 individuals in general population)(39). The *AMPD1* case with severe clinical symptoms also harboured homozygous pathogenic truncated variant in *CAPN3* which may result in the greater disease severity. Further studies are needed to resolve the gene contributions and for better understanding of AMPD1’s clinical significance. The diagnosis of myoadenylate deaminase deficiency is challenging given about 2% of muscle biopsies may have enzyme deficiency without clinical correlation (40-42). Deep phenotyping, immunohistochemical studies, western blot analysis or muscle imaging as well as functional studies are necessary to verify this group of myopathies.

The interesting finding of 13 patients with pathogenic variants in more than one myopathy gene (Table 5) and showing unusual presentations suggest a possible role of synergistic-heterozygosity and digenic contribution to unusual myopathies, similar to our recent finding in the US LGMD population (6). Multigenic combinations of pathogenic variants were detected involving 15 genes and 13 patients in this study suggest a high prevalence of synergistic heterozygosity in genetic myopathies. In some cases, the phenotype exhibited features of both the genes (eg: *GNE* and *MYOT*, patient #118; *DYSF* and *ANO5*, patient# 50, in Table 5) and in some patient’s phenotype favored one gene over the other (eg: patient# 125, Table 5). In some other case, there were clinical features unexplained by the identified genetic variants such as patient# 110 (Table 5) showing GNE-myopathy features without any *GNE* variant but pathogenic variants in *POMT1* and *CAPN3*. Thus, when faced with an atypical phenotype of inherited myopathy, the possibility of pathogenic variants in more than one myopathic gene exists and clinical exome or genome sequencing should be considered. Moreover, result of genetic testing in such multigenic cases need to be interpreted cautiously and muscle immunostaining should be considered. For multigenic myopathies, some cases may be due to genetically unidentified or a novel gene, especially those with heterozygous VUS in genes known to cause only recessive diseases. With widespread use of extensive panels, focused or whole-exome, and genome-sequencing, more such instances will be unearthed, helping further understanding of the pathophysiology and expressions of the genetic abnormalities. Functional omics platforms such as RNA-sequencing, proteomics, and metabolomics (43, 44) of the muscle tissue and segregation studies are needed in these unusual cohorts, to understand the variable expressions of the genes at the effector organ and the phenotypic variability. We propose to carry out this work in future which is a current limitation of this study.

True negative findings for the 14 patients with myopathy clinical symptoms suggest the need for whole genome sequencing (WGS) in the future to discover new myopathy causal genes harboring variants such as large deletions or duplication or deep-intronic variants that may have been missed with ES. A study limitation was that even though our inclusion criteria did not have any age restriction for subject recruitment the neuromuscular clinic caters mainly to the adult patients and hence pediatric cases were underrepresented in this study. Though this is a single center study, patients were referred to the center from all over the Indian subcontinent with diverse ethnicities and religious origins. Another limitation of our study was the lack of segregation analysis of compound heterozygous variant combinations identified in patients of our cohort. This is due to difficulty in the current setting in the region based on from where the patients travel and come to clinic and unavailability of parents or children of the patients. Segregation study is needed that will further enhance our understanding of the variant distribution in the population and for better family planning.

The paucity of population-based genetic testing in Indian subcontinent in public databases which may lead to insufficiency of minor allele frequency, lack of reports of similar myopathy patient(s) harboring the variant or unavailable functional data is a limitation in this study. Yet, this also points towards the immediate need and importance of studies such as ours on genetic testing with clinical correlation on large Asian cohorts, either disease-specific or general healthy population.

Finally, to our knowledge, the current study is the first comprehensive clinical-exome sequencing effort on a large genetic myopathy cohort in the Indian subcontinent that has enhanced our understanding of the spectrum of gene-variant-myopathy-subtype landscape in India yielding a high diagnostic rate. In this genomic era, studies such as this in different developing countries and continents on specific disease cohorts with diverse ethnicities will enable enhancement of the repertoire in the global genomic databases such as ExAc (http://exac.broadinstitute.org/), gnomAD (https://gnomad.broadinstitute.org/) and also country-specific large databases such as the UK100,000 Genome project (https://www.genomicsengland.co.uk/about-genomics-england/the-100000-genomes-project/), NIH All of US Genomics program (https://allofus.nih.gov/) and others for faster and more accurate variant classification, faster enhanced diagnostics, and the understanding of genotype-phenotype correlations.

## Data Availability

The original contributions presented in the study are included in the article/supplementary material; further inquiries can be directed to the corresponding author/s. This data can be found here: https://doi.org/10.5061/dryad.tmpg4f4w6

## AUTHOR CONTRIBUTIONS

SC and MH had full access to all data in this study and take responsibility for the integrity of the data and the accuracy of the data analysis. SC and MH designed, conceptualized and oversaw the entire study. Acquisition, analysis, or interpretation of data was done by SC, BRRN, SK, MS, AB, RD, PG, LR, and LG. Drafting of the manuscript was done by SC, SK, MS. Critical revision of the manuscript for important intellectual content was done by all authors. Statistical analysis was performed by SC, BRRN, SK, RD, PG, and LR. SC obtained funding. SC, MH and SK supervised the entire study.

## ACKNOWLDGEMENTS

The authors thank the patients and their families for study participation, and the funding sources.

## SUPPLEMENTARY MATERIAL

### Supplementary Methods

**Questionnaires for patient recruitment**

Table S1. Clinical symptoms and gene diagnosis of molecularly diagnosed patients

